# Language model–assisted label refinement for accurate sepsis detection from electronic health records

**DOI:** 10.64898/2026.09.08.26362428

**Authors:** Ioannis Kitsos Kalyvianakis, Claudio Pérez de Amézaga, Eun Sun Lee, Ann Palmer, Joseph Garner, Ulysses Shawdee Wu, Agni Orfanoudaki

## Abstract

Sepsis is a leading cause of hospital mortality, yet timely recognition is hampered by non-specific presentations and label noise in code-based case definitions. We developed STRIDE, a machine-learning framework for sepsis detection across seven hospitals with a scalable approach to label quality. We used a large language model applied to discharge summaries to refine encounter-level labels, with 349 physician-adjudicated encounters informing prompt development and serving as a clinical reference for evaluation. We compared 8-, 24-, and 48-hour observation windows and benchmarked against SOFA, SIRS, and Epic. Among 352,415 eligible encounters, STRIDE-8 achieved ROC-AUCs of 0.949 in derivation testing and 0.871 in physician-adjudicated evaluation. Removing SIRS-related predictors reduced these values to 0.853 and 0.743. Llama reclassified 12.8% of initially positive derivation encounters as nonseptic. These results support further investigation of short-window physiological screening combined with retrospective label review and evaluation against independent clinical reference outcomes.

## Introduction

Sepsis is a life-threatening medical condition that arises when the body’s response to an infection becomes dysregulated, resulting in widespread inflammation, tissue damage, and potential organ dysfunction [1]. It remains one of the leading causes of mortality in hospital settings, where delayed diagnosis can significantly impact patient outcomes. Each year, approximately 1.7 million adults in the United States develop sepsis, and around 270,000 of them ultimately die as a result [2]. Early intervention is crucial for survival, but the nonspecific and evolving presentation of sepsis makes reliable recognition difficult, often leading to delayed recognition and treatment initiation [3].

Traditional screening tools, such as the Systemic Inflammatory Response Syndrome (SIRS) criteria [4] and the Sequential Organ Failure Assessment (SOFA) score [5], together with guideline-based early recognition and management strategies [6], have long served as foundational methods for sepsis identification. SIRS offers high sensitivity by flagging a broad range of inflammatory responses. However, it is not specific to sepsis: many patients who meet SIRS criteria have no infection, so the criteria have poor specificity and a high false-positive rate, which can contribute to alert fatigue [7]. In contrast, SOFA, though more specific, relies on lab values and organ dysfunction markers that manifest later in the disease course, limiting its utility for early detection and itself is an organ dysfunction score which is not necessarily diagnostic of sepsis. More recently, commercial systems such as the Epic Sepsis Model have been implemented in electronic health record systems, promising automated and real-time risk alerts. However, studies have raised concerns about the transparency and generalizability of proprietary models like Epic, which in some settings have shown substantially suboptimal performance compared to the one reported by its developer [8].

These established screening instruments illustrate the core tension between sensitivity and specificity. High false positive rates in many early warning systems can increase unnecessary clinical evaluations, contribute to resource strain, and promote clinician alert fatigue [8, 9]. Conversely, making the alerting criteria more conservative to suppress false positives comes at the cost of more false negatives, risking missed cases and delayed intervention when timely treatment is essential [10]. This trade-off is especially important because sepsis becomes easier to detect as it progresses, as organ dysfunction and clear clinical signs appear, but the effectiveness of treatment diminishes with each hour of delay [3, 11].

A less visible but equally consequential obstacle lies in the labels used to build and benchmark these tools. Studies show that coded sepsis rates have risen sharply without matching clinical evidence [12–14]. Rhee et al. 2015 compared data from two academic hospitals between 2003 and 2012 and found a dramatic increase in sepsis International Classification of Diseases (ICD) codes (up to +706% for severe sepsis) while objective markers like positive blood cultures, shock, or elevated lactate remained stable or declined [12]. This illustrates how coding for sepsis has become more inclusive, extending the label to patients with less definitive evidence of severe infection. Rhee et al. (2014) illustrated that sepsis-coded hospitalizations increased while hospitalizations coded for common infections remained stable or declined. They also noted declining reported sepsis mortality, suggesting that changes in coding practices may partly explain these trends [15]. In addition, evolving definitions of sepsis have caused inconsistencies in documentation that contribute to overdiagnosis [13,14]. Any model trained on billing codes therefore risks learning to reproduce documentation behavior rather than physiology.

Machine learning has nonetheless advanced sepsis recognition considerably, and much of this work has explicitly moved away from billing codes toward clinical definitions. COMPOSER issues alerts while abstaining on cases it deems out-of-distribution, reducing false alarms [16]; the Risk of Sepsis score, trained on objective clinical-surveillance criteria rather than diagnostic codes, reports AUCs of 0.93–0.97 [17]; and the Sepsis ImmunoScore, validated against a physician-adjudicated clinical reference standard, became the first FDA-authorized AI tool for sepsis, with AUCs of 0.85 in derivation and 0.81 in external validation [18]. Effective feature engineering and temporal modeling have proven critical for handling high-dimensional, noisy, and irregularly sampled electronic health record (EHR) data: gradient-boosted trees over dynamic vital-sign features yield robust predictions [19], while recurrent neural network-based models such as DeepAISE capture evolving risk through time [20]. More recent work has extended this line of research with deep-learning and probabilistic approaches, including validation across multiple sites and international settings [21–24].

Yet important limitations persist even in these more advanced approaches. First, label quality remains a fundamental constraint: whether labels come from billing codes or from clinical consensus definitions, they are generated automatically from structured proxies (suspicion of infection inferred from culture and antibiotic timing, organ dysfunction inferred from score changes) and are rarely validated against the clinical record at scale. While some approaches have incorporated unstructured clinical notes to improve sepsis identification [25, 26] or used physician adjudication to validate labels [18], these strategies are rarely combined and seldom applied at the scale of a full derivation cohort, leaving residual label noise that propagates into both training and evaluation. Second, many models commit to a single fixed observation window, leaving open how much patient history is actually needed to characterize a patient’s current state at the point of care. Together, these gaps motivate the framework we present here.

To address these gaps, we developed STRIDE (Sepsis detection through Text-based Refinement of Information from Diagnostic Evidence), a machine-learning framework for detecting sepsis in the current 8-hour interval using EHR data from seven hospitals in the Hartford HealthCare system. Our approach treats label quality as a primary challenge to address. To this end, we determine the sepsis labels based on a combination of diagnosis codes, clinical criteria, and infection-related orders and then refine them further using unstructured text. Specifically, to achieve this at scale, we use the large language model (LLM) Llama 3.1-8B Instruct over discharge summaries to reclassify encounters inconsistent with sepsis. Physician adjudication of 349 initially positive encounters informed development of the LLM prompt and provided a separate clinical reference for evaluating STRIDE. Moreover, rather than fixing a single observation window, we ask how much preceding patient history is needed to characterize a patient’s current sepsis state, comparing models that use the most recent 8-, 24-, and 48-hour windows. We frame the task as contemporaneous detection rather than onset forecasting, benchmark against the tools clinicians routinely use and assess calibration alongside discrimination. An overview of the study design, including the label-refinement, feature-extraction, and model-derivation steps, is shown in Fig. 1.

**Figure 1.**
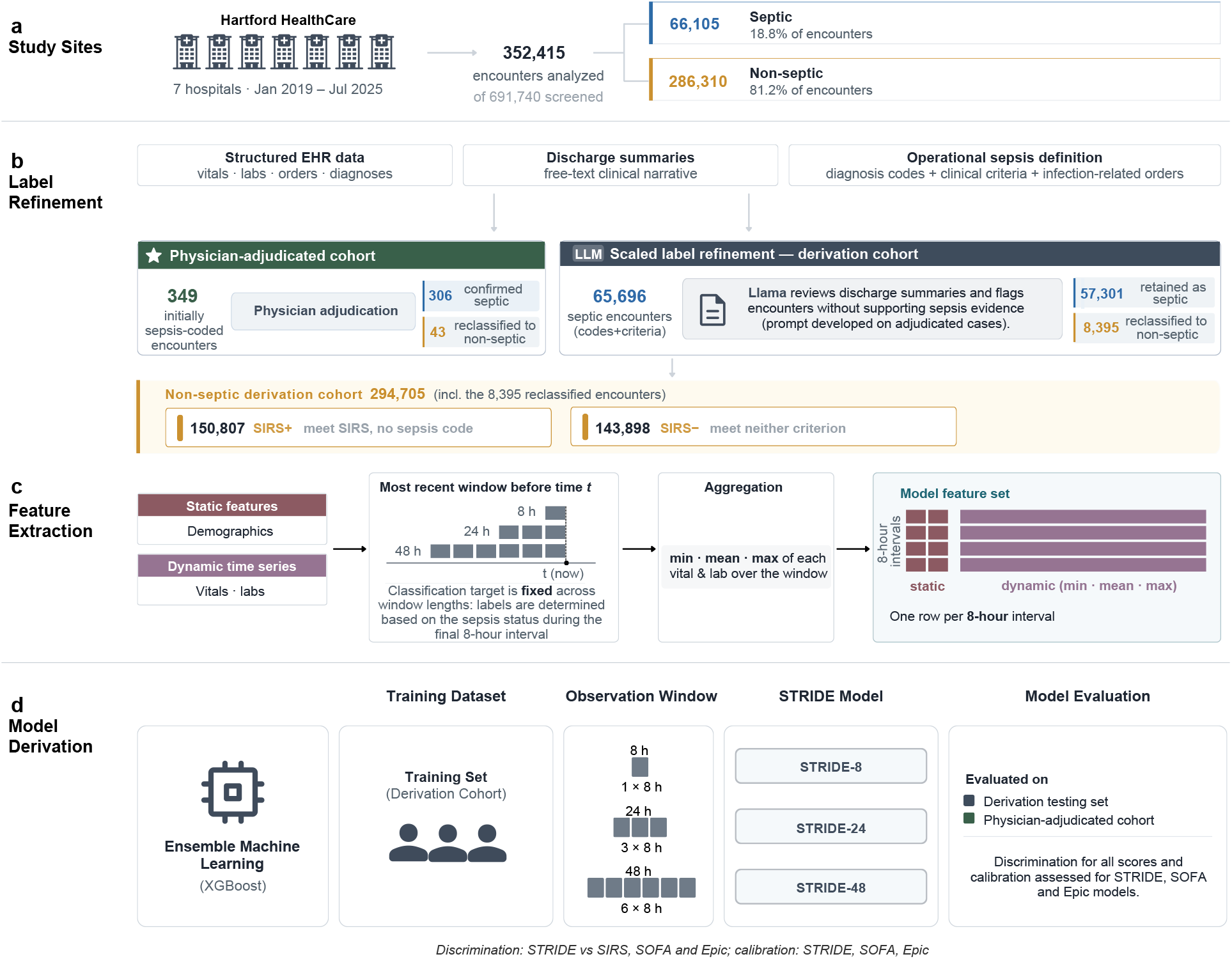
Overview of the STRIDE study design. **a**, Study sites and cohort definition; **b**, Label refinement; **c**, Feature extraction; **d**, Model derivation. Icons: Healthicons (CC0 1.0, public domain) and Google Material Symbols (Apache 2.0).

## Results

### Study Population: Inclusion and Exclusion Criteria

We initially identified 77,394 encounters with a sepsis diagnosis code and 614,346 non-septic encounters from January 2019 to July 2025, across seven hospitals in the Hartford HealthCare system. After applying our predefined exclusion criteria, including high missingness (*>*40% missing features), insufficient length of stay (*<*48 hours of data), discontinuous time series and the absence of clinical criteria for sepsis in septic encounters, a total of 352,415 encounters (66,105 initially septic and 286,310 non-septic) remained for analysis. Fig. 2 illustrates this cohort selection process in detail. Excluded sepsis-coded encounters had shorter stays and higher in-hospital mortality than included ones (14.6% versus 11.5%; Supplementary Table S15). More information about the exclusion criteria can be found in Methods.

**Figure 2.**
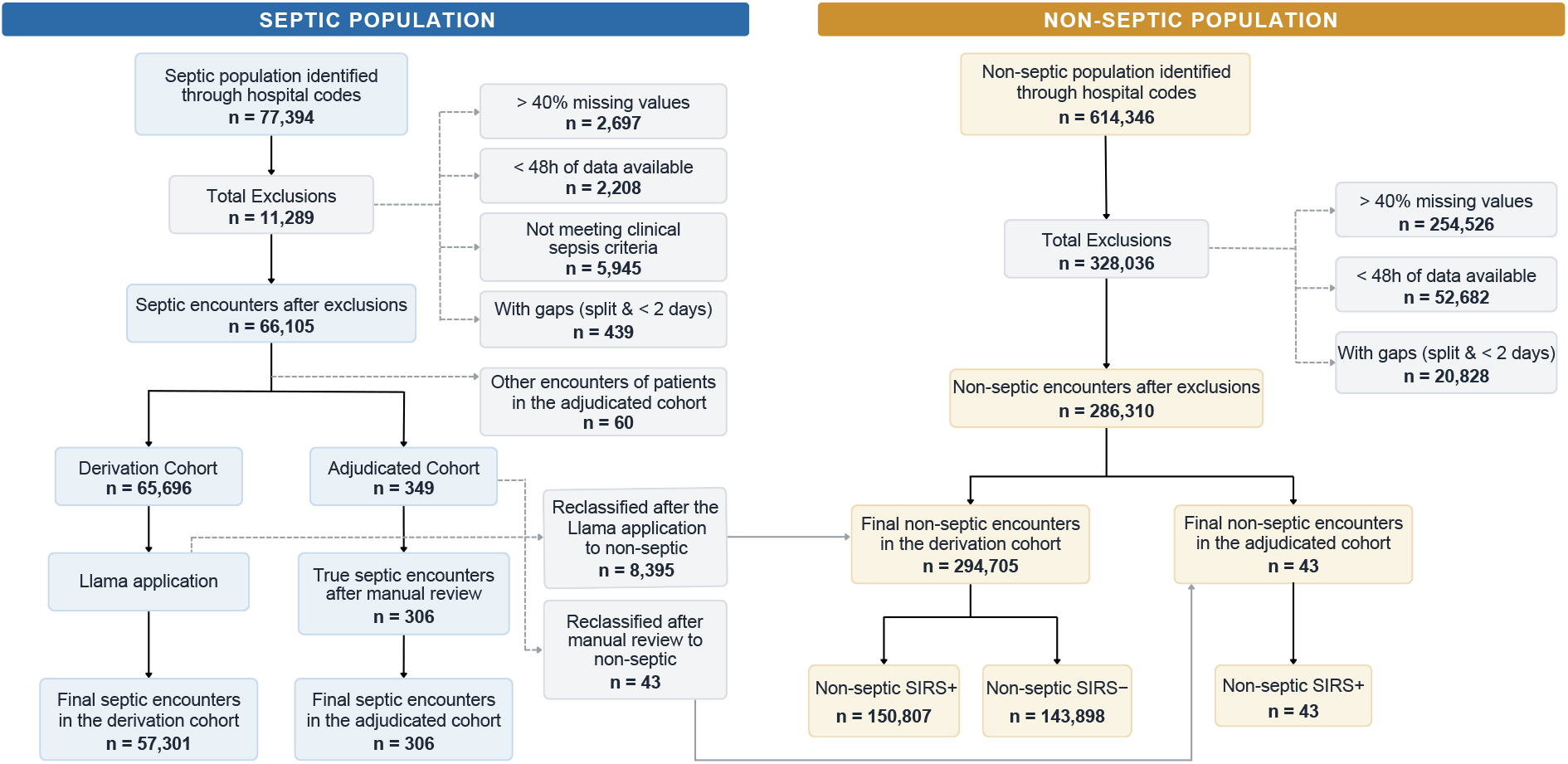
Patient cohort selection flowchart. Systematic selection of septic and nonseptic encounters from Hartford HealthCare system (January 2019–July 2025) after applying predefined exclusion criteria.

### Study Population: Decomposition of Septic and Non-septic Encounters

The 66,105 initially septic encounters were divided into a physician-adjudicated cohort and a derivation cohort (Fig. 1b). We randomly selected 349 encounters for physician review. To keep the two cohorts patient-disjoint, we also excluded from derivation 60 further sepsiscoded encounters belonging to 46 patients in the adjudicated cohort, leaving 65,696 initially positive derivation encounters. Infectious disease specialists confirmed sepsis in 306 of the 349 adjudicated encounters and reclassified 43 as non-septic. These physician labels were used to develop the LLM prompt and to evaluate STRIDE.

The septic derivation cohort underwent processing using the Llama model (detailed in Language Model-Based Label Refinement), which reclassified 8,395 of the 65,696 initially positive derivation encounters as non-septic. The 57,301 encounters (41,950 unique patients) that retained a positive label after Llama review formed the septic derivation cohort; all had a sepsis-related diagnosis code and met the operational clinical criteria.

The non-septic population in derivation included the initially non-septic encounters and those reclassified by Llama. The final derivation cohort comprised 352,006 encounters from 224,756 patients, divided into 150,807 non-septic SIRS+ encounters (113,713 patients) and 143,898 non-septic SIRS− encounters (101,739 patients), alongside the septic cohort. More details about the cohorts can be found in Methods.

### Study Population: Cohort Characteristics

The encounter characteristics of the derivation and physician-adjudicated cohort are summarized in Table 1.

**Table 1:** Descriptive characteristics of the study population at the encounter level, calculated using mean values aggregated within 8-hour intervals. Vital signs and laboratory measurements are encounter means of 8-hour interval means, before multivariable imputation.

|  | Derivation Cohort |  |  | Physician-adjudicated Cohort |  |
|---|---|---|---|---|---|
| **Variable** | **Septic** | **Non-septic SIRS+** | **Non-septic SIRS−** | **Septic** | **Non-septic SIRS+** |
| **Population** |  |  |  |  |  |
| Number of Patients | 41,950 | 113,713 | 101,739 | 306 | 43 |
| Number of Encounters | 57,301 | 150,807 | 143,898 | 306 | 43 |
| **Demographics** |  |  |  |  |  |
| Age at admission, mean (SD) | 67.1 (16.8) | 62.6 (19.7) | 63.3 (19.5) | 67.9 (16.9) | 60.3 (19.0) |
| Men, n (%) | 30,093 (52.5%) | 73,029 (48.4%) | 65,900 (45.8%) | 166 (54.2%) | 21 (48.8%) |
| Women, n (%) | 26,984 (47.1%) | 77,241 (51.2%) | 77,383 (53.8%) | 137 (44.8%) | 21 (48.8%) |
| Unknown, n (%) | 224 (0.4%) | 537 (0.4%) | 615 (0.4%) | 3 (1.0%) | 1 (2.3%) |
| **Race, n (%)** |  |  |  |  |  |
| White or Caucasian | 42,910 (74.9%) | 108,102 (71.7%) | 103,942 (72.2%) | 234 (76.5%) | 29 (67.4%) |
| Other | 8,620 (15.0%) | 23,709 (15.7%) | 22,127 (15.4%) | 34 (11.1%) | 7 (16.3%) |
| Black or African American | 5,029 (8.8%) | 16,531 (11.0%) | 15,782 (11.0%) | 32 (10.5%) | 6 (14.0%) |
| Unknown | 742 (1.3%) | 2,465 (1.6%) | 2,047 (1.4%) | 6 (2.0%) | 1 (2.3%) |
| **Ethnicity, n (%)** |  |  |  |  |  |
| Not Hispanic or Latino | 48,564 (84.8%) | 126,183 (83.7%) | 120,856 (84.0%) | 268 (87.6%) | 33 (76.7%) |
| Hispanic or Latino | 7,831 (13.7%) | 21,598 (14.3%) | 20,547 (14.3%) | 33 (10.8%) | 7 (16.3%) |
| Unknown | 906 (1.6%) | 3,026 (2.0%) | 2,495 (1.7%) | 5 (1.6%) | 3 (7.0%) |
| **Smoking status, n (%)** |  |  |  |  |  |
| Never smoker | 21,260 (37.1%) | 57,936 (38.4%) | 59,140 (41.1%) | 116 (37.9%) | 14 (32.6%) |
| Former smoker | 21,674 (37.8%) | 48,593 (32.2%) | 45,802 (31.8%) | 102 (33.3%) | 16 (37.2%) |
| Current smoker | 7,794 (13.6%) | 21,778 (14.4%) | 21,866 (15.2%) | 49 (16.0%) | 6 (14.0%) |
| Never assessed | 1,780 (3.1%) | 6,763 (4.5%) | 4,676 (3.2%) | 10 (3.3%) | 2 (4.7%) |
| Other / unknown | 4,793 (8.4%) | 15,737 (10.4%) | 12,414 (8.6%) | 29 (9.5%) | 5 (11.6%) |
| **Admission type, n (%)** |  |  |  |  |  |
| Emergency | 52,963 (92.4%) | 113,221 (75.1%) | 114,949 (79.9%) | 270 (88.2%) | 33 (76.7%) |
| Urgent | 2,911 (5.1%) | 9,353 (6.2%) | 9,854 (6.8%) | 21 (6.9%) | 8 (18.6%) |
| Elective | 817 (1.4%) | 21,040 (14.0%) | 16,748 (11.6%) | 8 (2.6%) | 0 (0.0%) |
| Other | 610 (1.1%) | 7,193 (4.8%) | 2,347 (1.6%) | 7 (2.3%) | 2 (4.7%) |
| **Length of stay (days), n (%)** |  |  |  |  |  |
| 0–2 | 5,478 (9.6%) | 27,246 (18.1%) | 45,927 (31.9%) | 21 (6.9%) | 3 (7.0%) |
| 3–5 | 21,277 (37.1%) | 63,317 (42.0%) | 63,454 (44.1%) | 94 (30.7%) | 8 (18.6%) |
| 6–8 | 12,269 (21.4%) | 28,027 (18.6%) | 18,653 (13.0%) | 82 (26.8%) | 15 (34.9%) |
| 9+ | 18,277 (31.9%) | 32,217 (21.4%) | 15,864 (11.0%) | 109 (35.6%) | 17 (39.5%) |
| **Vital Signs, mean (SD)** |  |  |  |  |  |
| Heart rate (bpm) | 85.5 (11.8) | 83.2 (12.5) | 75.1 (10.6) | 85.5 (11.7) | 89.0 (14.5) |
| Respiratory rate (breaths/min) | 19.2 (2.3) | 18.8 (3.1) | 17.8 (1.1) | 19.5 (2.5) | 19.8 (2.4) |
| Systolic BP (mmHg) | 125.3 (14.6) | 127.6 (15.5) | 130.9 (15.8) | 123.5 (14.8) | 123.0 (13.7) |
| Diastolic BP (mmHg) | 69.0 (7.7) | 70.9 (8.6) | 72.0 (8.3) | 68.1 (8.1) | 68.4 (8.7) |
| Temperature (°F) | 98.3 (0.7) | 98.0 (0.7) | 97.9 (0.5) | 98.4 (0.8) | 98.3 (0.8) |
| Oxygen saturation (%) | 95.9 (1.7) | 96.2 (1.7) | 96.6 (1.6) | 95.9 (1.6) | 96.3 (1.6) |
| **Lab Results, mean (SD)** |  |  |  |  |  |
| Bilirubin (mg/dL) | 0.8 (1.8) | 0.9 (1.9) | 0.7 (1.1) | 1.0 (1.7) | 1.9 (5.1) |
| Alanine Aminotransferase (U/L) | 32.5 (29.0) | 33.2 (29.8) | 28.6 (26.9) | 35.1 (29.5) | 39.5 (33.9) |
| Aspartate Aminotransferase (U/L) | 38.4 (29.9) | 39.4 (31.3) | 32.7 (26.6) | 42.7 (31.7) | 51.0 (36.7) |
| Alkaline phosphatase (U/L) | 112.6 (61.7) | 103.2 (56.0) | 95.1 (49.8) | 110.5 (60.1) | 123.0 (70.5) |
| Blood Urea Nitrogen (mg/dL) | 25.4 (19.1) | 21.0 (16.0) | 18.4 (13.4) | 28.3 (22.4) | 24.2 (22.6) |
| Creatinine (mg/dL) | 1.3 (1.3) | 1.2 (1.2) | 1.1 (1.1) | 1.4 (1.3) | 1.2 (0.9) |
| Platelets (×10<sup>9</sup>/L) | 238.3 (114.1) | 235.6 (98.7) | 232.3 (84.7) | 214.6 (107.7) | 236.9 (118.4) |
| Potassium (mmol/L) | 4.0 (0.4) | 4.1 (0.4) | 4.0 (0.4) | 4.0 (0.5) | 4.0 (0.3) |
| Sodium (mmol/L) | 138.5 (3.8) | 138.1 (3.4) | 138.5 (3.2) | 138.7 (4.2) | 138.5 (4.5) |
| Chloride (mmol/L) | 103.1 (5.0) | 102.3 (4.5) | 102.9 (4.1) | 103.3 (5.4) | 103.6 (4.9) |
| Albumin (g/dL) | 3.1 (1.1) | 3.5 (0.6) | 3.8 (0.5) | 3.1 (0.6) | 3.1 (0.6) |
| White Blood Cell Count (×10<sup>9</sup>/L) | 11.5 (6.1) | 10.2 (5.4) | 7.8 (2.9) | 11.6 (5.6) | 11.2 (4.8) |
| CO<sub>2</sub> (mmol/L) | 24.5 (3.9) | 24.9 (3.7) | 25.1 (3.2) | 24.5 (3.7) | 23.5 (2.9) |
| **Other, mean (SD)** |  |  |  |  |  |
| Height (cm) | 168.1 (11.4) | 167.0 (15.4) | 167.7 (11.3) | 167.4 (11.8) | 169.7 (11.5) |
| Weight (kg) | 83.8 (27.0) | 82.4 (25.0) | 81.9 (23.1) | 83.6 (24.1) | 81.9 (25.4) |

### Language Model-Based Label Refinement

To enhance label precision across the large derivation cohort, we integrated a language-model-based label refinement step (Fig. 3). We applied Llama 3.1-8B Instruct [27] to analyze discharge summaries and classify septic encounters as truly septic or non-septic. In the 349 physician-adjudicated encounters used to develop the prompt, Llama agreed with physician labels in 314 cases (90.0%; 95% CI 86.4–92.7%). Sensitivity was 93.8% and specificity was 62.8% (Supplementary Table S12). Of the encounters Llama retained, physicians had confirmed sepsis in 94.7% (287/303), compared with 87.7% (306/349) of all sampled encounters. We then applied this model across the septic-labeled encounters in the derivation cohort. As a result, 8,395 encounters (12.8%) were reclassified by Llama as non-septic. Patients with any reclassified encounter were assigned only to training, so the test sets in the main analysis contained no reclassified encounters; Supplementary Table S13 evaluates alternative allocations. Further details about the use of Llama are provided in the Methods.

**Figure 3.**
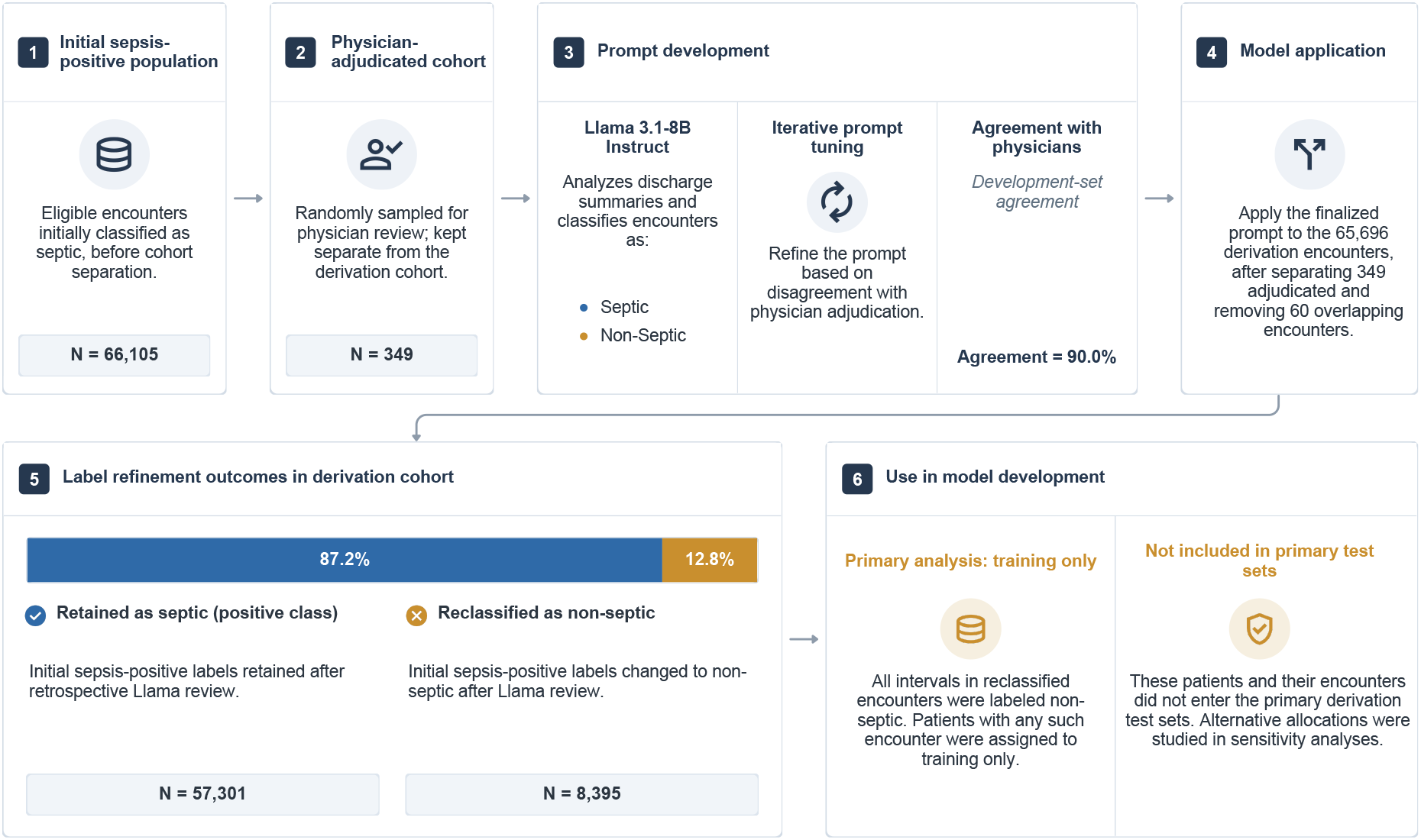
Workflow for language model-based label refinement. Icons: Google Material Symbols (Apache 2.0).

Demographics and length of stay were analyzed per encounter. Septic encounters involved older individuals (mean age 67.1 years, SD 16.8) compared to non-septic SIRS+ (62.6 years, SD 19.7) and SIRS− groups (63.3 years, SD 19.5). Men represented 52.5% of septic, 48.4% of SIRS+, and 45.8% of SIRS− encounters. Prolonged hospitalizations (≥9 days) occurred in 31.9% of septic encounters versus 21.4% for non-septic SIRS+ and 11.0% for non-septic SIRS−. Only 9.6% of septic encounters resulted in discharge within 0–2 days (vs. 31.9% in non-septic SIRS−). The physician-adjudicated cohort mirrored these trends. More details about the clinical features can be found in Table 1 and Supplementary Table S14.

### Predictive Model Performance

We evaluated three gradient-boosted (XGBoost) models, hereafter STRIDE-8, STRIDE-24, and STRIDE-48, that differ in how much preceding patient history they use to classify the patient’s current sepsis state. STRIDE-8 uses only the most recent 8-hour interval, a single static snapshot with no prior context, whereas STRIDE-24 and STRIDE-48 incorporate the three and six consecutive 8-hour intervals leading up to the detection point, respectively, allowing them to capture progressively longer-term physiological trends (full specification in Methods, Table 7). Using these windows, we assessed both discrimination and calibration of the models on the testing set of the derivation cohort. STRIDE-8 and STRIDE-24 achieved mean ROC-AUCs of 0.949, followed by STRIDE-48 at 0.945; corresponding ROC-AUCs in the physician-adjudicated cohort were 0.871, 0.870 and 0.861, respectively (Table 2).

**Table 2:** Discrimination of STRIDE across observation windows in the derivation test cohort and the physician-adjudicated cohort. Values are means over five patient-level splits with 95% confidence intervals from a patient-level bootstrap. AP represents the average precision (area under the precision–recall curve). The no-skill value of AP equals the proportion of positive intervals (5.8% in derivation testing, 22.9% in the physician-adjudicated cohort), so AP is not comparable between cohorts.

| Model | Test ROC-AUC | Adjudicated ROC-AUC | Test AP | Adjudicated AP |
|---|---|---|---|---|
| STRIDE-8 | 0.949 (0.947–0.950) | 0.871 (0.828–0.906) | 0.458 (0.445–0.470) | 0.557 (0.447–0.655) |
| STRIDE-24 | 0.949 (0.948–0.951) | 0.870 (0.830–0.903) | 0.477 (0.466–0.488) | 0.566 (0.463–0.658) |
| STRIDE-48 | 0.945 (0.943–0.947) | 0.861 (0.819–0.896) | 0.423 (0.408–0.438) | 0.512 (0.414–0.614) |

To assess performance across different populations, we further evaluated all models across multiple encounter subpopulations of the derivation cohort, including an only septic encounter cohort, and populations where non-septic encounters either met or did not meet SIRS criteria. These stratifications included both septic and non-septic encounters, with the SIRS+ and SIRS− subgroups differentiated based on the SIRS status of the non-septic patients. As shown in Table 3, STRIDE-8 consistently outperformed SOFA and Epic across evaluation groups and retained high discrimination when non-septic encounters meeting SIRS criteria were included alongside septic encounters.

**Table 3:** ROC-AUC with 95% confidence intervals (patient-level bootstrap) for sepsis detection methods using an 8-hour time window. Analyses use interval-level labels. The septic subset includes positive and negative intervals from septic encounters; the mixed groups additionally include intervals from their respective non-septic encounter groups.

| **Method** | **Testing set of derivation cohort** | **Septic subset of derivation cohort** | **Septic + non-septic SIRS+ encounters** | **Septic + non-septic SIRS– encounters** | **Physician-adjudicated cohort** |
|---|---|---|---|---|---|
| STRIDE-8 | 0.949<br>[0.947–0.950] | 0.907<br>[0.903–0.911] | 0.926<br>[0.924–0.928] | 0.963<br>[0.961–0.965] | 0.871<br>[0.828–0.906] |
| SOFA | 0.742<br>[0.736–0.747] | 0.665<br>[0.657–0.673] | 0.706<br>[0.699–0.712] | 0.761<br>[0.755–0.766] | 0.637<br>[0.585–0.682] |
| Epic | 0.828<br>[0.822–0.834] | 0.725<br>[0.718–0.732] | 0.789<br>[0.782–0.795] | 0.842<br>[0.836–0.848] | 0.724<br>[0.677–0.766] |
| SIRS | 0.891<br>[0.890–0.893] | 0.878<br>[0.874–0.882] | 0.849<br>[0.847–0.852] | 0.942<br>[0.940–0.944] | 0.848<br>[0.813–0.879] |

STRIDE-8 demonstrated robust performance across all cohorts, achieving a ROC-AUC of 0.907 [0.903–0.911] in the septic subgroup, 0.926 [0.924–0.928] in the SIRS+ population, and 0.963 [0.961–0.965] in the SIRS− population. The latter two comparisons add non-septic encounters to the septic subset; both therefore contain positive and negative intervals. Among the clinical comparators, SIRS achieved the highest discrimination (0.891 in derivation testing and 0.848 in the adjudicated cohort), followed by Epic (0.828 and 0.724; evaluated on intervals with a recorded score) and SOFA (0.742 and 0.637). STRIDE-8 exceeded all three comparators in every evaluation group, although its confidence interval overlapped that of SIRS in the adjudicated cohort. Because SIRS is part of the operational outcome definition, its sensitivity is 100% by construction, and its ROC-AUC reflects a single operating point of a binary rule rather than a ranking of risk. Performance metrics are reported as means across five patient-level splits, with 95% confidence intervals from patient-level bootstrapping. Fig. 4a illustrates the ROC curves comparing STRIDE-8 with clinical benchmarks on both the standard testing set and the physician-adjudicated cohort.

**Figure 4.**
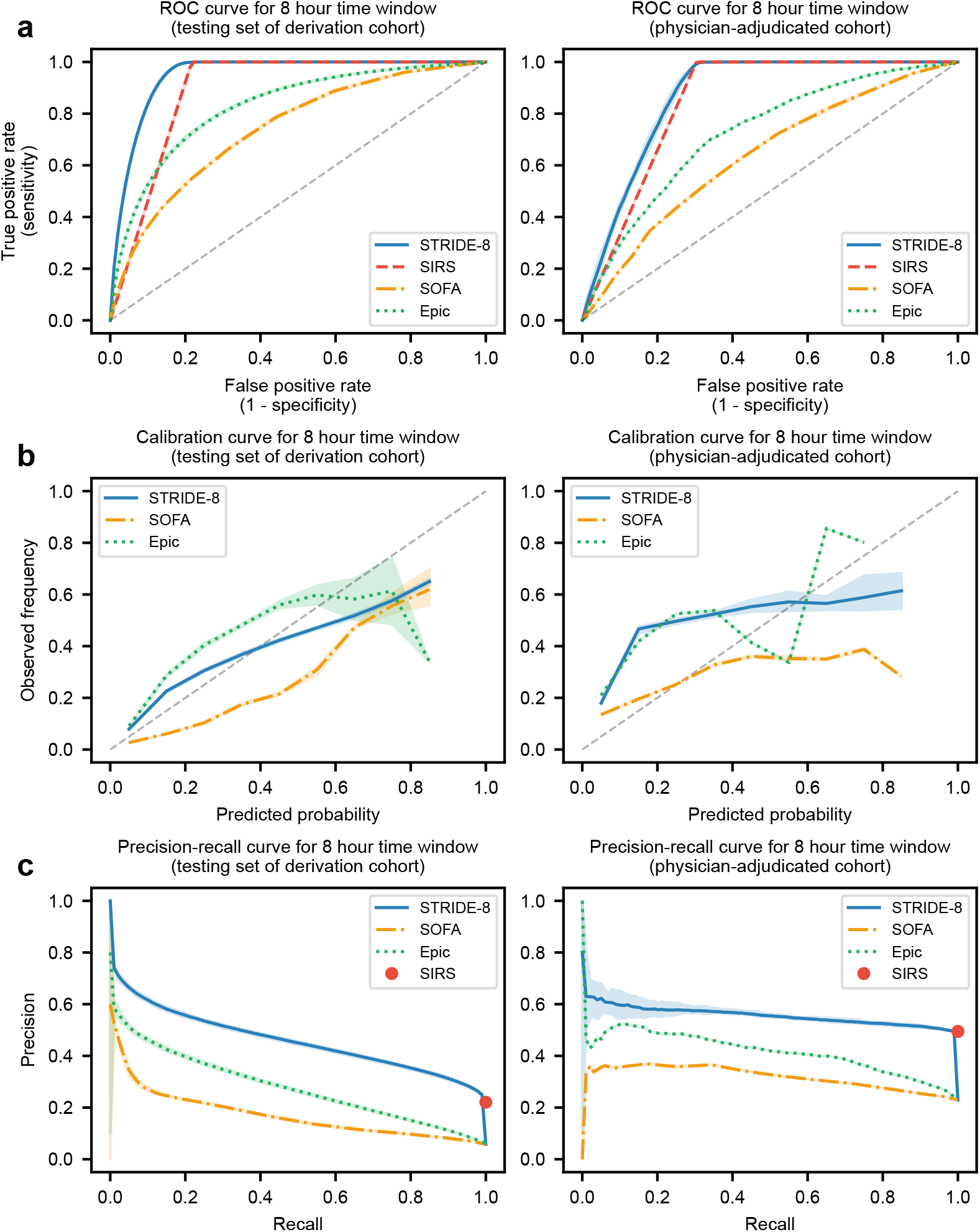
Discrimination and calibration of STRIDE-8 against clinical scores for the 8-hour window. Receiver operating characteristic (**a**), calibration (**b**), and precision-recall (**c**) curves for STRIDE-8 and the clinical benchmarks (SOFA, Epic, and SIRS) on the testing set of the derivation cohort (left) and the physician-adjudicated cohort (right). Solid lines show the mean across the five patient-level splits and shaded bands the spread across splits; the dashed diagonal denotes chance (**a**) or perfect calibration (**b**). SIRS is shown as a red circle (operating point) in **c** and is omitted from **b**. Calibration panels show predicted probabilities for STRIDE and min–max-normalized scores for Epic and SOFA.

To further assess model reliability, we evaluated calibration and precision-recall performance across all time windows for the testing set of the derivation cohort and the physician-adjudicated cohort. For the 8-hour window (Fig. 4b), STRIDE-8 achieved an expected calibration error (ECE) of 0.015 and a Brier score of 0.040 in the testing set. In the physician-adjudicated cohort, the corresponding ECE and Brier score were 0.105 and 0.143. Full precision-recall and calibration metrics for all windows are reported in Supplementary Table S4.

The 8-hour window precision-recall analysis showed that STRIDE-8 had higher average precision (0.458 in derivation testing and 0.557 in the physician-adjudicated cohort) than Epic (0.277 in derivation testing) and SOFA (0.159 in derivation testing) (Fig. 4c). Because SIRS is a fixed binary rule, we represent it as a point in Fig. 4c and do not compute an area-based summary for it. Detailed results for the 24-hour and 48-hour windows are available in Supplementary Tables S1 and S4–S7 and Supplementary Figs. S1–S3.

To further evaluate the model’s clinical applicability, Table 4 summarizes specificity and accuracy at a target sensitivity of 80% for the 8-hour window model in comparison with clinical screening tools. We focused on STRIDE-8 because extending the observation window to 24 or 48 hours did not improve discrimination over the most recent 8-hour window, indicating that, for contemporaneous sepsis detection, recent physiological status was sufficient and additional historical context provided limited incremental value.

**Table 4:** Operating-point performance for the 8-hour window. Thresholds were selected on derivation data to target 80% sensitivity; SIRS is evaluated at its fixed rule. Values are means over five patient-level splits with 95% confidence intervals (patient-level bootstrap). Counts are true positives (TP), false positives (FP), true negatives (TN) and false negatives (FN) of 8-hour intervals, means per split. Epic was evaluated only on intervals with a recorded score. Epic thresholds are shown on the min–max normalized scale, with the corresponding recorded Epic Sepsis Model score in parentheses (normalized value = recorded score/93.13, the maximum recorded score; minimum 0).

| **Method** | **Cohort** | **Threshold** | **Sensitivity** | **Specificity** | **PPV** | **NPV** | **Accuracy** | **TP / FP**<br>**TN / FN** |
|---|---|---|---|---|---|---|---|---|
| STRIDE-8 | Testing set of<br>derivation cohort | 11.4% | 80.0%<br>[79.4%–80.6%] | 91.0%<br>[90.7%–91.2%] | 35.3%<br>[34.2%–36.3%] | 98.7%<br>[98.6%–98.7%] | 90.4%<br>[90.1%–90.6%] | 90,461 / 165,684<br>1,675,943 / 22,620 |
|  | Physician-adjudicated cohort | 11.4% | 80.9%<br>[77.6%–84.4%] | 78.1%<br>[72.3%–83.4%] | 52.4%<br>[44.6%–59.5%] | 93.2%<br>[91.5%–94.9%] | 78.8%<br>[74.2%–82.8%] | 2,284 / 2,075<br>7,419 / 539 |
| SOFA | Testing set of<br>derivation cohort | 2 points | 88.8%<br>[88.2%–89.5%] | 40.2%<br>[39.3%–41.1%] | 8.4%<br>[8.0%–8.7%] | 98.3%<br>[98.2%–98.4%] | 43.1%<br>[42.2%–43.9%] | 100,461 / 1,100,410<br>741,217 / 12,620 |
|  | Physician-adjudicated cohort | 2 points | 90.0%<br>[86.4%–93.0%] | 21.4%<br>[16.3%–27.0%] | 25.4%<br>[20.9%–30.1%] | 87.9%<br>[84.9%–90.4%] | 37.1%<br>[31.8%–42.2%] | 2,542 / 7,462<br>2,032 / 281 |
| Epic | Testing set of<br>derivation cohort | 3.2%<br>(3.01) | 80.0%<br>[79.0%–80.9%] | 70.6%<br>[70.1%–71.4%] | 15.0%<br>[14.5%–15.6%] | 98.2%<br>[98.1%–98.3%] | 71.2%<br>[70.7%–71.9%] | 84,574 / 477,400<br>1,146,706 / 21,131 |
|  | Physician-adjudicated cohort | 3.2%<br>(3.01) | 82.5%<br>[77.8%–86.6%] | 48.1%<br>[41.6%–54.5%] | 32.9%<br>[27.5%–38.3%] | 89.9%<br>[86.8%–92.4%] | 56.2%<br>[51.1%–60.9%] | 2,124 / 4,329<br>4,008 / 451 |
| SIRS | Testing set of<br>derivation cohort | ≥2 criteria | 100.0%<br>[100.0%–100.0%] | 78.3%<br>[78.0%–78.6%] | 22.1%<br>[21.3%–22.8%] | 100.0%<br>[100.0%–100.0%] | 79.6%<br>[79.2%–79.9%] | 113,081 / 399,673<br>1,441,955 / 0 |
|  | Physician-adjudicated cohort | ≥2 criteria | 100.0%<br>[100.0%–100.0%] | 69.6%<br>[62.7%–75.8%] | 49.5%<br>[42.6%–55.5%] | 100.0%<br>[100.0%–100.0%] | 76.6%<br>[70.8%–81.0%] | 2,823 / 2,884<br>6,610 / 0 |

STRIDE-8 achieved a specificity of 91.0% [90.7%–91.2%] and an accuracy of 90.4% [90.1%– 90.6%] in the derivation testing set, and a specificity of 78.1% [72.3%–83.4%] with an accuracy of 78.8% [74.2%–82.8%] in the physician-adjudicated cohort. Compared with STRIDE-8, SOFA showed substantially lower specificity (40.2% in derivation testing; 21.4% in the adjudicated cohort), while Epic reached a specificity of 70.6% and 48.1%, respectively. At its fixed decision rule, SIRS sensitivity was 100% in both cohorts, with specificity of 78.3% and 69.6%; its high sensitivity reflects the inclusion of SIRS in the operational label. The different methods were not necessarily evaluated at identical sensitivities (Table 4).

### SIRS-related predictors and subgroup performance

To examine the contribution of SIRS-related predictors, we retrained STRIDE without heart rate, respiratory rate, temperature and white blood cell count. STRIDE-8 ROC-AUC decreased from 0.949 to 0.853 in derivation testing and from 0.871 to 0.743 in the physician-adjudicated cohort. Without these variables, STRIDE-24 achieved ROC-AUCs of 0.868 and 0.751, and STRIDE-48 achieved 0.867 and 0.745, respectively. We used the same patient partitions and outcome labels as in the main analysis. We also evaluated model performance among intervals meeting at least two SIRS criteria. Once SIRS was present, an interval received a positive label only if the encounter had a sepsis diagnosis code and a culture or imaging order was assigned to that interval. STRIDE-8 achieved ROC-AUCs of 0.765 (0.759–0.770) in derivation testing and 0.576 (0.515–0.631) in the physician-adjudicated cohort, compared with 0.685 and 0.568 for Epic (Table 5). Discrimination was lower for both models in this restricted analysis.

**Table 5:** ROC-AUC of the full and SIRS-feature-ablated 8-hour STRIDE models and clinical comparators. The ablation uses the same outcome labels and patient partitions as the main analysis. The last two columns are restricted to intervals meeting at least two SIRS criteria; Epic values use intervals with a recorded score. The ablated model was not evaluated in this subset.

|  | All intervals |  | Intervals meeting ≥2 SIRS criteria |  |
|---|---|---|---|---|
| Model | Derivation test | Physician-adjudicated | Derivation test | Physician-adjudicated |
| STRIDE-8, full model | 0.949 | 0.871 | 0.765 | 0.576 |
| STRIDE-8, without SIRS features | 0.853 | 0.743 | – | – |
| Epic sepsis score | 0.828 | 0.724 | 0.685 | 0.568 |
| SOFA | 0.742 | 0.637 | 0.632 | 0.513 |

For STRIDE-8 without SIRS-related features, ROC-AUC was 0.755 in the septic subset, 0.819 when non-septic SIRS-positive encounters were added, and 0.861 when non-septic SIRS-negative encounters were added (Supplementary Table S17). STRIDE-8 ROC-AUC ranged from 0.938 to 0.967 across hospitals and from 0.943 to 0.952 across calendar years (Supplementary Table S11).

### Operating thresholds and alert burden

At 80% sensitivity in derivation testing, STRIDE-8 yielded a positive predictive value of 35.3% and a negative predictive value of 98.7%, with 5.8% prevalence of positive intervals. The thresholds selected in derivation were then applied to the physician-adjudicated cohort, yielding 80.9% sensitivity, 78.1% specificity, 52.4% positive predictive value and 93.2% negative predictive value. At the same operating point, STRIDE-8 produced 0.39 positive classifications per patient-day in derivation testing, including 0.25 false positives. These corresponded to 10.6 false-positive intervals per 1,000 observed patient-hours. At least one positive interval occurred in 93.6% of septic and 26.6% of non-septic encounters; 85% of positive intervals followed an earlier positive interval within the same encounter. Consecutive positive intervals were counted separately.

### Feature importance

Fig. 5 displays the 15 most influential features for each time window (8, 24, and 48 hours), with features ordered in the plots according to their mean absolute SHapley Additive exPlanations (SHAP) value [28].

**Figure 5.**
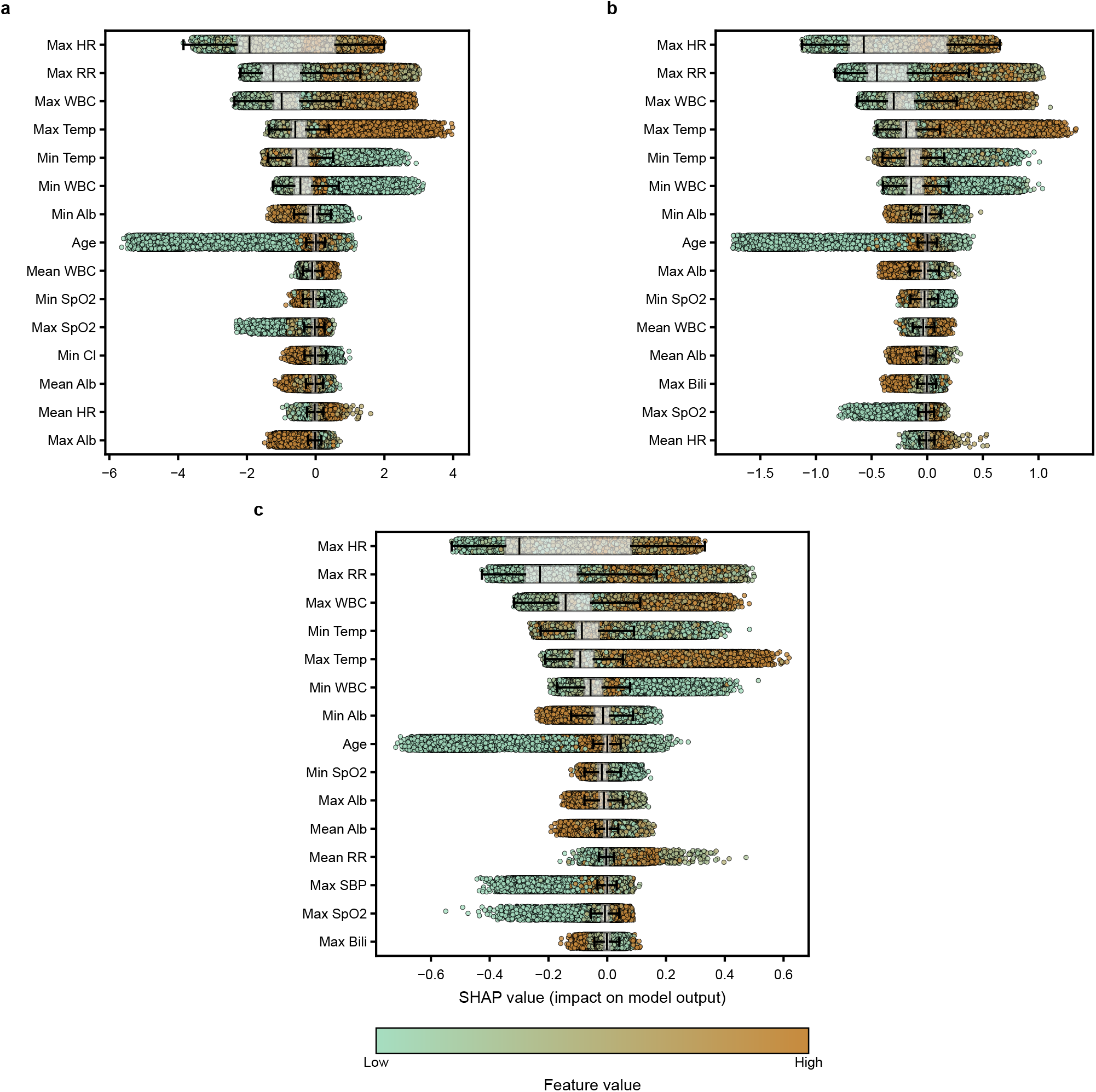
Feature importance analysis using SHAP values for STRIDE-8, STRIDE-24, and STRIDE-48. SHAP values for the 15 highest-ranked features of the models fitted on the first patient-level partition, computed on its test set, for the 8-hour (**a**), 24-hour (**b**) and 48-hour (**c**) windows. Each dot is one observation window; for multi-interval windows, SHAP values are averaged over the 8-hour intervals in the window. Positive values push the output towards sepsis. Dot color shows the feature value and boxplots summarize the SHAP distribution. Features are ordered by mean absolute SHAP value; x-axis ranges differ between panels. HR, heart rate; RR, respiratory rate; WBC, white blood cell count; Temp, temperature; Alb, albumin; SpO_2_, oxygen saturation; Cl, chloride; Bili, bilirubin; SBP, systolic blood pressure.

For the 8-hour window, the SHAP analysis revealed that maximum heart rate was the most influential predictor, followed by maximum respiratory rate and maximum white blood cell count. Maximum temperature ranked fourth in importance, while minimum temperature was the fifth most important feature. Minimum white blood cell count, minimum albumin, age, mean white blood cell count and minimum oxygen saturation also ranked among the most influential features.

In the 24-hour model, maximum heart rate similarly emerged as the top predictor, with maximum respiratory rate and maximum white blood cell count following as the second and third most important features. Temperature, white blood cell count, albumin, age and oxygen saturation were also influential across the 24- and 48-hour models.

Across all windows, maximum heart rate, maximum respiratory rate, and maximum white blood cell count consistently emerged as the top predictors of model output. Temperature-related features (maximum and minimum) also ranked highly in importance. This pattern is expected because heart rate, respiratory rate, white blood cell count, and temperature are components of the SIRS criteria, which form part of the operational sepsis label used in this study. The models also use continuous summaries of these variables and additional measurements, such as albumin and oxygen saturation. Further details on the feature importance analysis are provided in the Methods.

Table 6 compares the key features driving our model’s detections with the variables used in established clinical scoring systems and prior sepsis literature, revealing varying degrees of feature overlap and divergence. STRIDE-8 identified maximum heart rate, maximum respiratory rate, and maximum white blood cell count as the most influential predictors, followed by maximum temperature and minimum temperature. Several of these variables also appear in the SIRS criteria, while other high-ranking predictors, including albumin, age and oxygen saturation, extend beyond them. The Sepsis ImmunoScore [18] prioritized procalcitonin as its top feature, a biomarker not available in our dataset, though it shared respiratory rate, temperature, and albumin with our feature set. Similarly, the Risk of Sepsis Score [17] emphasized maximum lactic acid as its leading feature, which was not used in our model, though both models identified maximum white blood cell count, respiratory rate, and temperature as important features.

**Table 6:**
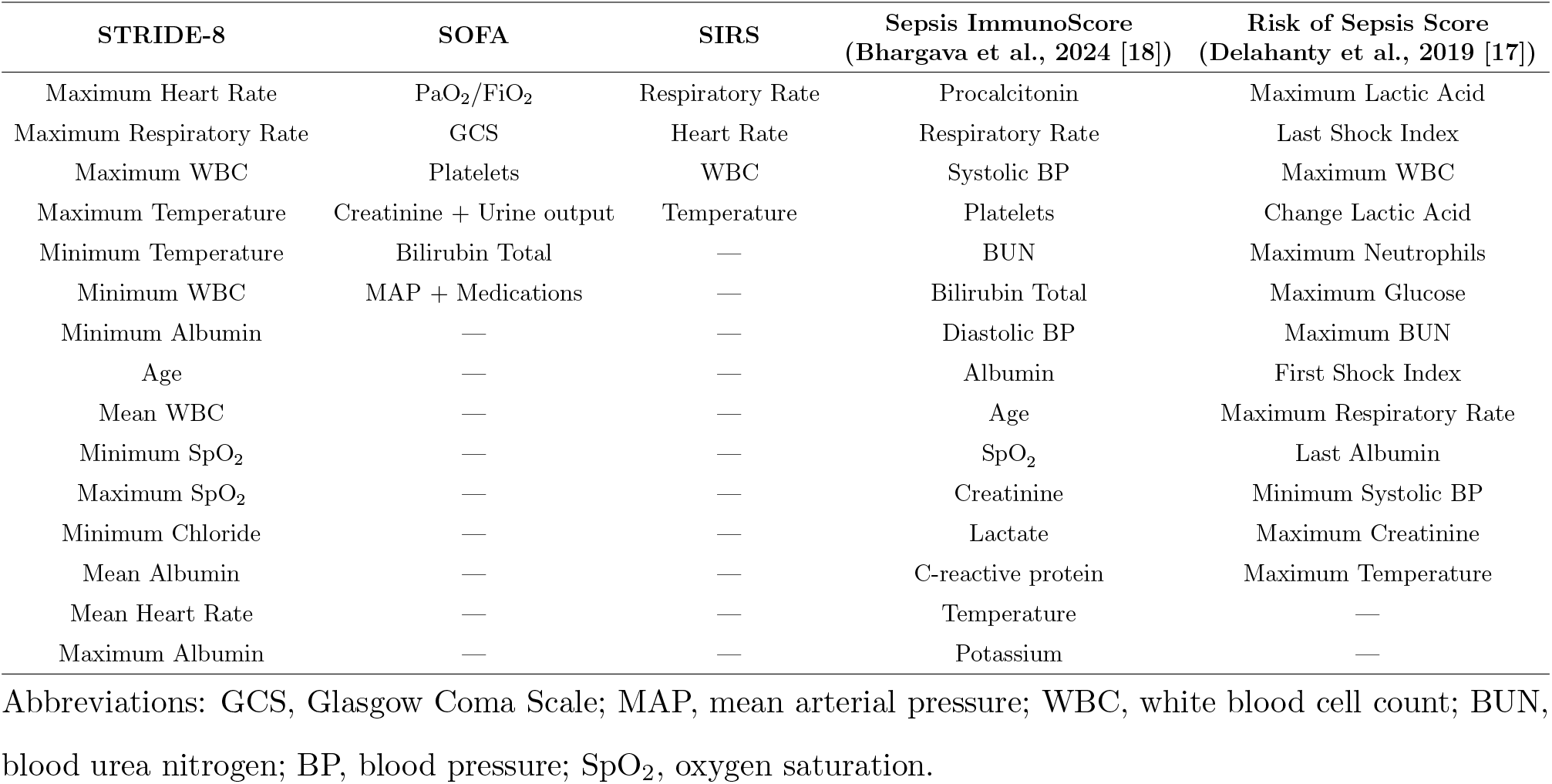
Feature comparison: the 15 highest-ranked STRIDE-8 features by mean absolute SHAP value (Fig. 5a) versus the inputs of clinical and published sepsis scores.

**Table 7:**
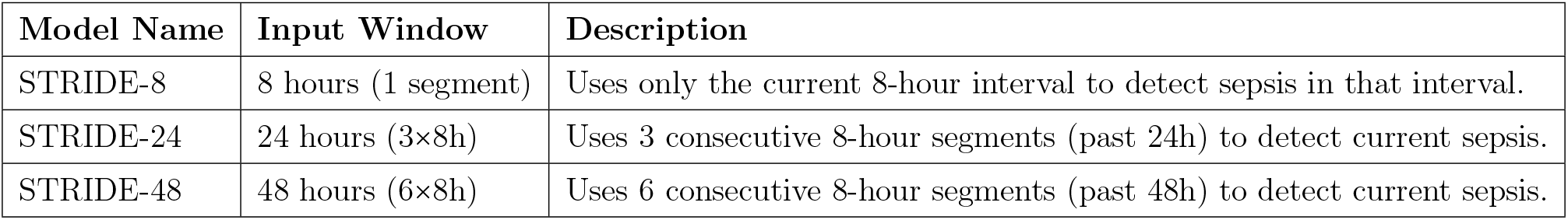
Sepsis detection models by length of observation window.

| **Model Name** | **Input Window** | **Description** |
|---|---|---|
| STRIDE-8 | 8 hours (1 segment) | Uses only the current 8-hour interval to detect sepsis in that interval. |
| STRIDE-24 | 24 hours (3×8h) | Uses 3 consecutive 8-hour segments (past 24h) to detect current sepsis. |
| STRIDE-48 | 48 hours (6×8h) | Uses 6 consecutive 8-hour segments (past 48h) to detect current sepsis. |

## Discussion

We developed and evaluated STRIDE, a machine-learning framework for dynamic sepsis detection using vital signs and laboratory measurements from seven Hartford HealthCare hospitals. We first refined encounter labels using discharge summaries and then trained the detection models on structured clinical measurements. Three observations emerge from the analyses. First, the label used for training is itself a design decision with measurable consequences. Second, recent physiology carries most of the information needed to characterize a patient’s current status. Third, the performance of any model evaluated against an operational definition is bounded by what that definition encodes.

Broad operational definitions and administrative codes are known to overflag sepsis [29], while unstructured clinical notes can help refine case identification [25,26,30]. Our adjudication illustrated the first point directly, as specialists rejected sepsis in roughly one in eight sampled encounters that carried both a sepsis code and the operational clinical criteria. A language model applied to discharge narratives makes it possible to audit labels across an entire derivation cohort, a scale that manual review cannot reach. We used physician-reviewed encounters to develop the Llama prompt and then applied it to discharge summaries in the derivation cohort. In the prompt-development cases, physicians confirmed sepsis more often among encounters Llama retained than among coded encounters overall (94.7% versus 87.7%), but they also confirmed sepsis in 19 of 46 encounters that Llama classified as non-septic (Supplementary Table S12). Label refinement is therefore a trade-off on the labels themselves rather than a correction, and the balance chosen should be stated explicitly. The prompt also asked whether an infection was documented, not whether a sepsis definition was met. It therefore targets sepsis codes assigned to patients without a documented infection, rather than misjudged severity or organ dysfunction. STRIDE performed similarly in the adjudicated cohort however the reclassified encounters were allocated (Supplementary Table S13). In this study, the main contribution of refinement therefore lies in the composition and auditability of the training labels. Establishing whether it improves detection requires a reference standard independent of both the diagnosis codes and the prompt.

In the eligible encounters, all of which had at least 48 hours of recorded data, longer input windows added little to discrimination. Part of this follows from the task. Because each label describes the current interval, the most recent measurements are the most directly informative, and earlier trajectories add little to characterizing present status. Longer histories may matter more for forecasting onset, which this study did not address. For contemporaneous classification, a short window has practical advantages. It requires less preceding data, is less exposed to gaps in the record, and can begin classifying once 8 hours of data are available rather than 24 or 48. To capture the variability of frequently measured variables within each interval, the model summarizes every 8-hour period by its minimum, maximum and mean. This design preserves fluctuations that a single reading could miss, as in other temporal representations of clinical time series [31, 32].

The most influential predictors were SIRS components, which is expected because SIRS forms part of the outcome definition. This pattern also aligns with systematic reviews showing that dynamic vital signs and laboratory values are the most influential features in machine learning models for sepsis prediction [33]. Two analyses clarify what this overlap means. Without the SIRS measurements, STRIDE still discriminated better than Epic and SOFA in both cohorts, drawing on variables such as albumin, oxygen saturation and age (Table 5). The model therefore uses information beyond the SIRS criteria. Among intervals already meeting SIRS criteria, however, discrimination fell for every method, and the remaining variation in the label depended on sepsis coding and the presence of culture or imaging orders. These reflect clinical practice as well as the patient’s condition, and the lower discrimination is consistent with the difficulty of recovering order-dependent labels from physiological measurements. This illustrates a broader point. Operational definitions built from orders and codes encode clinician behavior, and models trained and evaluated against them partly learn that behavior. Under such labels, the ceiling on physiology-based detection is set by the label as much as by the model.

The comparison with established tools should be read in light of what each measures. SIRS performs well against this outcome largely because it is part of it. SOFA quantifies organ dysfunction, which typically appears later and is not specific to infection. The Epic Sepsis Model was developed under its vendor’s own outcome definition and alerting configuration, which we could not reproduce [8]. Lower discrimination for these tools against a contemporaneous, SIRS-based label does not show how they would perform against other clinical definitions. Rather, the comparison shows how much of the operational target structured physiology can recover beyond simple rules.

Interval classifications are not alerts. Most positive classifications followed an earlier positive interval in the same encounter, so the burden on clinicians would depend less on per-interval specificity than on the alerting policy built around the model, including how repeated positives are consolidated, when alerts are suppressed after acknowledgement, and which patients are monitored. Assessing STRIDE as a clinical tool requires specifying such a policy and evaluating it prospectively. This study did not assess whether STRIDE shortens time to treatment or improves patient outcomes.

Several limitations should be considered. Our data come from a single health system, and although it spans seven hospitals, these share an electronic health record and common clinical practices, so external validation across independent systems is needed before broader deployment. The operational sepsis definition combined SIRS, clinical orders used as a proxy for infection investigation, and sepsis diagnosis codes. The order criterion included all entries in the imaging table, extending beyond studies ordered specifically for suspected infection. Physicians established whether sepsis occurred during an encounter but did not assign independent onset times. Diagnosis codes were also visible to them, and paired ratings were not retained. The adjudicated cohort contained only initially sepsis-positive encounters and also informed prompt development, so it is neither an independent test of Llama nor a basis for estimating performance among patients without a sepsis code. Performance was examined by hospital, calendar year and demographic group, but not by ward or illness severity, and calibration was not assessed within subgroups.

Label refinement also remains imperfect. Refinement was applied only to the positive class, so sepsis missed by coding remains among the negatives. The septic cohort was identified from billing codes and infection-related criteria, whose accuracy depends on documentation quality, and the language model relies on clinical notes that are themselves subjective and written after the hospitalization. These narratives influenced only the training labels and were not available to STRIDE as predictors. Future work could incorporate additional automated tools or manual chart review to strengthen labels in both classes.

Finally, incomplete and irregularly sampled data required imputation and aggregation, which may introduce bias, particularly because interpolation can draw on later observations. Omitting predictor interpolation reduced but did not remove discrimination (Supplementary Table S10). Prospective use would nonetheless require a fully time-causal preprocessing pipeline and an independently timed clinical reference. The eligibility criteria also excluded short stays and encounters with substantial missingness, and excluded sepsis-coded encounters had higher in-hospital mortality than included ones, so STRIDE’s performance in these patients is unknown (Supplementary Table S15). Some informative variables, most notably procalcitonin, were not routinely measured and could not be included [34, 35].

Our results support combining retrospective review of coded sepsis labels with short-window physiological detection, and indicate that the reference outcome, rather than the model, is the main constraint on evaluating this approach. Independent, time-specific clinical adjudication that includes coded-negative encounters would allow both stages to be evaluated without overlap between predictors and outcome definition, and would make it possible to measure when STRIDE classifications occur relative to clinical recognition. Together with prospective assessment of a defined alerting policy, it is the most important next step in evaluating the clinical use of STRIDE.

## Methods

### Exclusion Criteria

We began by identifying hospital encounters labeled as septic or non-septic using diagnosis codes. The EHR data covered January 2019 to July 2025 across seven Hartford HealthCare hospitals and included diagnosis codes, clinical orders, demographic characteristics, vital signs and laboratory results. To ensure high data quality and temporal continuity for modeling, several exclusion criteria were applied. Encounters were eligible regardless of age, so the cohort includes some newborn encounters with neonatal sepsis codes (Supplementary Table S16); adult SIRS thresholds were applied to all encounters. The encounter inclusion diagram for both septic and non-septic encounters can be seen in Fig. 2. Encounters with more than 40% missing feature values after initial interpolation were excluded, as high levels of missingness compromise the reliability of downstream analyses (see Table S2 of the Supplementary Information). We also excluded encounters with fewer than 48 hours of recorded data, so all three models could be evaluated on a common study population. After these preprocessing steps, we applied our previously defined operational sepsis criteria and the septic encounters that did not meet this definition were excluded. Finally, we required that the data for each encounter span consecutive calendar days; if gaps in the time series were present, the encounters were split and any resulting segment shorter than two days was excluded. Characteristics of included and excluded encounters are compared in Supplementary Table S15. Reporting was guided by TRIPOD+AI [36].

### Sepsis Definition and Label Refinement: Operational Definition of Sepsis

For the purposes of this study, we define sepsis using criteria established by the 2001 International Sepsis Definitions Conference [37], which operationalized sepsis as the presence of systemic inflammatory response syndrome (SIRS) in the context of suspected or confirmed infection. Specifically, a patient is considered septic during any 8-hour interval in which all of the following conditions are met:

1. SIRS criteria are satisfied [4]. This includes meeting at least two of the standard SIRS thresholds (See Section 3 of the Supplementary Information for further details).
2. At least one of the following records was assigned to the interval as an operational indicator of suspected infection:

a. A culture order (e.g., blood, urine, sputum)
b. An imaging-table record (e.g., radiograph or CT; the table also includes non-radiological studies).
3. The hospital encounter is associated with a sepsis-related diagnosis code.

This operational definition uses the 2001 consensus criteria [37] and combines physiological evidence of systemic inflammation with clinical investigation for a possible infection. Culture orders were assigned to intervals using their recorded time, while imaging records were assigned to all three intervals of the order date. The imaging criterion included all entries in the imaging table, regardless of body region or modality (Supplementary Section 5). Labels were generated using recorded and interpolated measurements before multivariable imputation, and each positive label referred to the current 8-hour interval.

### Sepsis Definition and Label Refinement: Llama 3.1-8B Instruct

Sepsis diagnosis codes in administrative data are known to have variable accuracy, often leading to false positives due to overcoding, retrospective documentation practices, or cases where sepsis is suspected but ultimately ruled out [15]. In our initial derivation cohort, we observed that a subset of encounters labeled as septic based on diagnosis codes and operational clinical criteria did not exhibit clinical features consistent with sepsis, raising concerns about label quality and potential noise in model training.

To address this issue, we implemented a label refinement strategy using the Llama 3.1-8B Instruct language model [27], which enabled integration of unstructured clinical narrative into the labeling process (Fig. 3). As described in the Physician-adjudicated Cohort Overview section, we used physician-adjudicated encounters reviewed by Infectious Disease specialists to develop the Llama prompt and provide clinical reference labels for STRIDE evaluation.

We developed the prompt through an iterative refinement process, adjusting it based on agreement with the physician-adjudicated labels. The clinical reviews were completed before prompt development, and the same cases were used to assess agreement with Llama; they were not an independent test of the prompt. To focus the analysis on the most clinically relevant text, the Llama model was provided with the ”Hospital Course” section from each discharge summary (or the full summary when that section could not be identified) and prompted to return a binary (yes/no) classification regarding documented evidence of infection or sepsis. This approach offered a consistent and scalable interpretation of clinical text to complement structured data.

We then applied the finalized prompt to the remaining initially positive derivation encounters. Encounters that the model reclassified as non-septic were relabeled accordingly. Patients with reclassified encounters were assigned to training in the primary model evaluation. Initially non-septic encounters were not reassessed. Discharge narratives were used for label refinement and were not included as STRIDE predictors. Further technical details about the Llama model and prompt structure are available in Section 8 of the Supplementary Information.

### Sepsis Definition and Label Refinement: Derivation Cohort Overview

The derivation cohort was analyzed in three mutually exclusive groups: septic, non-septic SIRS-positive and non-septic SIRS-negative. Initial classification depended on sepsis diagnosis codes and the operational clinical criteria, followed by Llama refinement of initially positive encounters.

The first group consists of 57,301 encounters with a sepsis diagnosis code and the operational criteria that retained a positive label after Llama refinement. These are considered septic and constitute the positive class for model training and evaluation. The second group comprises encounters that did not receive a sepsis diagnosis code but did meet the SIRS and infection-investigation components of the clinical sepsis criteria. It also includes the encounters reclassified by Llama. These encounters were classified as non-septic either because they lacked a sepsis diagnosis code or because they were reclassified by Llama following retrospective review of their discharge summaries. The third group contains encounters that neither carried a sepsis diagnosis code nor met the clinical sepsis criteria at any point during hospitalization. Together, the second and third groups form the negative class used for model training and evaluation. The final two non-septic groups can be partitioned between the SIRS-positive and the SIRS-negative encounters.

This structure ensures that the model is trained on a broad spectrum of clinical presentations, including coded sepsis, cases with sepsis-like features but no clinical diagnosis, and encounters without the specified clinical criteria. This allowed performance to be assessed using both SIRS-positive and SIRS-negative non-septic encounters.

### Sepsis Definition and Label Refinement: Physician-adjudicated Cohort Overview

Each of the physician-adjudicated cohort encounters represents a unique patient randomly selected from the initially sepsis-positive eligible encounters and set aside from the derivation cohort. Specifically, these encounters had a hospital sepsis diagnosis code assigned and met the clinical sepsis criteria at least once during hospitalization. Each case was manually reviewed on a case-by-case basis by Infectious Disease specialists from Hartford HealthCare. The infectious disease experts reviewed the clinical record, including diagnoses, medical history and discharge documentation, and assigned a single encounter-level sepsis label. Original diagnosis codes were visible, but paired independent ratings were not retained, so inter-rater agreement could not be calculated. The review did not determine the time of sepsis onset. As a result of this rigorous adjudication process, 306 encounters were confirmed as truly septic, while 43 were reclassified as non-septic, reflecting the clinical ambiguity that often surrounds sepsis diagnosis. In confirmed encounters, 8-hour labels followed the operational clinical criteria; in rejected encounters, all intervals were considered non-septic.

### Feature Engineering and Modeling Approach: Feature Selection

Data were collected using the EHR database of the Hartford HealthCare hospital system. The features included in this study were selected in collaboration with Infectious Disease specialists at Hartford HealthCare to ensure clinical relevance to the detection of sepsis. The final dataset comprised 25 clinical and demographic variables: 6 demographic variables, 6 vital signs, and 13 laboratory-based clinical measurements. The 19 time-varying variables were each summarized by three statistics, giving 63 model inputs per 8-hour interval once the six demographic attributes were added. Demographic variables, age, sex, race, multiracial identification, ethnicity and smoking status, were treated as static for each hospital encounter. Time-varying features were extracted from structured EHR and selected to reflect physiologic domains known to be disrupted in sepsis, including inflammation and organ dysfunction. Vital signs are central to clinical scoring systems such as SIRS and SOFA, while the laboratory features reflect renal and hepatic function, inflammatory response and metabolic status. These measurements are routinely collected in inpatient settings and have demonstrated relevance to sepsis pathophysiology and clinical outcomes [38–41]. All features were selected for their clinical relevance, consistency across patient records, and utility in sepsis risk stratification. The interval timestamp was retained as a non-clinical temporal indexing variable associated with each 8-hour interval, used to preserve temporal ordering when constructing the observation windows; it was not a predictor, nor were patient identifiers or medical-history variables.

### Feature Engineering and Modeling Approach: Data Restructuring and Sliding Window

To prepare the data for modeling, time-varying features were aggregated at the observation level using fixed 8-hour intervals across each patient encounter. As a result, each day consists of three 8-hour intervals. Within each interval, we computed the mean, minimum, and maximum values for every continuous feature, resulting in a structured time-series representation of the patient’s physiological state. This approach enables the model to capture clinically meaningful trends in vital signs and laboratory results. After initial interpolation, each segment was evaluated for sepsis using our operational sepsis definition. A binary label was assigned to each interval: 1 if the patient met sepsis criteria during that interval, and 0 otherwise.

Missing values were initially addressed through linear interpolation, as many laboratory values used in the model are not recorded at consistent intervals, such as every 8 hours. Linear interpolation estimates missing values by assuming a straight-line relationship between adjacent data points. This approach helped to align the data and fill gaps in the time series, serving as an initial step in addressing missing values in our data set. Because interpolation used adjacent observations, it could include measurements recorded after the interval being classified. After linear interpolation, IterativeImputer was used to impute any remaining missing values [42]. For each split, an imputer fitted to pooled training observations from both classes was applied without refitting to test and adjudicated patients. Fitting used at most 200,000 sampled training rows, ten iterations and a split-specific random seed. A separate training-fitted imputer completed the additional measurements required to calculate SOFA. The interval labels had already been assigned and were not changed by this multivariable imputation. More details about the missing data imputation process can be found in Section 6 of the Supplementary Information.

To handle the temporal nature of the data, we applied a sliding window technique to transform the time-series data into a format suitable for supervised learning. This approach captures the temporal evolution of the variables by creating fixed-size windows of data that move sequentially across the time series with a specified shift interval. Each window moved forward by one 8-hour interval. For our analysis, we utilized three window configurations:

1. A window size of 6, where each window encompasses six consecutive 8-hour intervals, corresponding to a total duration of 48 hours.
2. A window size of 3, where each window encompasses three consecutive 8-hour intervals, corresponding to a total duration of 24 hours.
3. A window size of 1, where each window contains data from a single 8-hour interval.

For each multi-interval window, the target label was derived exclusively from the final interval in the sequence, enabling the model to learn from preceding physiological patterns to classify the patient’s current sepsis state. The single-interval model does not incorporate historical information; the label corresponds directly to the single 8-hour interval being analyzed. This design allows the models to leverage different depths of historical context while maintaining a consistent goal: identifying the patient’s sepsis status in the present. An overview of these configurations is presented in Table 7, and a detailed explanation of the sliding window approach is shown in Fig. 6 and Section 9 of the Supplementary Information. The 8-, 24- and 48-hour models used 63, 189 and 378 inputs, respectively.

**Figure 6.**
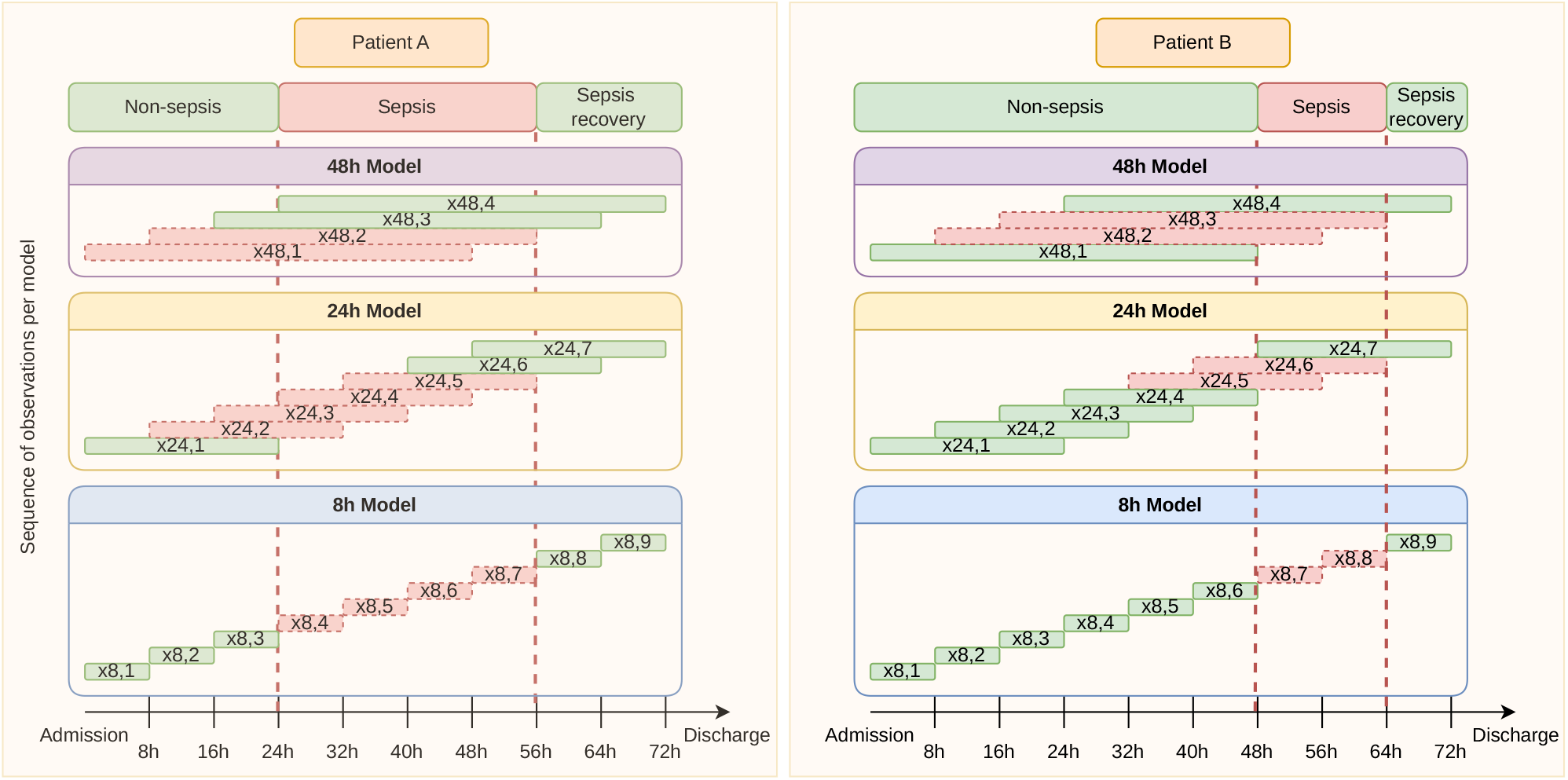
Representation of the different windows generated by the sliding window approach for the 8h, 24h, and 48h models. The length of each window corresponds to the time interval used to construct the predictor variables. A window is labeled as sepsis when its final 8-hour interval meets the operational criteria. Each observation *X^I^_m,t_* represents inpatient *I*, model *m* ∈ {8, 24, 48}, and time *t* ∈ {8, 16, 24*, . . .* }.

### Feature Engineering and Modeling Approach: Model Development

We trained XGBoost binary classification models to classify the presence of sepsis, with the outcome labeled as 1 if the patient had sepsis and 0 otherwise [43]. XGBoost performed similarly to Random Forest with an 8-hour window and slightly better with longer windows (Supplementary Section 12). Model training was performed using five repeated patient-level train–test splits of the full dataset. In each partition, the dataset was randomly split into training and testing sets, with approximately 70% of eligible patients assigned to the training set and the remaining patients to the test set. To maintain independence and prevent data leakage, all encounters from the same patient were assigned exclusively to either the training or testing subset within each split. Patients with any Llama-reclassified encounter were assigned to training in the primary analysis. The physician-adjudicated patients remained outside the model-fitting process. Supplementary Table S13 examines two alternative ways of allocating the reclassified encounters. Internal evaluation was conducted across the five randomized splits. For each split, performance metrics were calculated on the test set. These metrics were aggregated across all five splits, and both the average values and 95% confidence intervals were computed using patient-level bootstrap resampling within each split. Finally, model performance was evaluated on the physician-adjudicated cohort, which remained separate from STRIDE training.

For each model, StandardScaler was fitted on the training features and applied unchanged to evaluation patients. XGBoost used 100 estimators, maximum depth 12, random state=0, n jobs=64 and no class reweighting (package default); the other settings were also kept at their defaults (XGBoost 3.0.5, scikit-learn 1.7.2). The same specification was used across splits, without using test or physician-adjudicated labels to select hyperparameters. No hyperparameter search was performed, and the Random Forest comparison used the same values (Supplementary Section 12).

### Performance Evaluation: Model Evaluation

We evaluated model performance across three observation windows: 48 hours, 24 hours, and 8 hours, each ending at the interval being classified. Key metrics included the area under the receiver operating characteristic curve (ROC-AUC) and average precision from the precision-recall curve, which were chosen to capture both overall discrimination and performance in the presence of class imbalance. Calibration curves were generated for each time window to assess the agreement between predicted probabilities and observed outcome frequencies, providing insight into the reliability of the model’s risk estimates. To evaluate model performance at a clinically relevant operating point, we identified the probability threshold that achieved a target sensitivity of 80% using derivation test patients. Each derivation test set was divided into two groups of patients: a threshold was selected on one group and evaluated on the other, and then the groups were exchanged. The threshold selected from the full derivation test set was applied unchanged to the physician-adjudicated cohort. For the integer-valued SOFA score, the highest threshold achieving at least the target sensitivity was selected. SIRS was evaluated at its prespecified fixed rule. All performance metrics were reported as the mean and the corresponding 95% confidence intervals, calculated from 200 patient-level bootstrap resamples per split, pooled across the five splits. This approach ensured a robust assessment of model variability and generalization. To contextualize our results, we compared the predictive performance of our models to established clinical benchmarks, including the SOFA score, the Epic score and the SIRS.

SIRS was calculated from temperature, heart rate, respiratory rate and white blood cell count using the same recorded and interpolated measurements used to assign the interval labels. Missing components counted as not met. SOFA was computed from the available respiratory, coagulation, liver, cardiovascular, neurological and renal measures using the organ-specific scoring rules and the adaptations detailed in Supplementary Section 4.

Epic scores were obtained directly from the EHR and averaged within each 8-hour interval. They were normalized using the minimum and maximum of all recorded scores (0 and 93.13), and the thresholds are reported in both the recorded and normalized units. The deployed Epic model version was not available in the study records. The analysis of Epic was restricted to intervals with a recorded score. For SOFA and Epic, calibration summaries used min–max-normalized scores rather than calibrated event probabilities.

We calculated sensitivity, specificity, positive and negative predictive values, accuracy, F1 score, and the corresponding counts of true-positive, false-positive, true-negative and false-negative intervals. Calibration was summarized using the Brier score and expected calibration error (ECE) over ten equal-width bins. Positive interval classifications were also expressed per patient-day, without grouping consecutive positive intervals into alert episodes.

To examine the contribution of SIRS-related predictors, we retrained each model after excluding the minimum, mean and maximum values of temperature, heart rate, respiratory rate and white blood cell count. Patient partitions, outcome labels and model settings were unchanged. We also evaluated STRIDE-8, Epic and SOFA among intervals already meeting at least two SIRS criteria. Additional analyses examined performance by hospital, calendar year and recorded demographic characteristics, and sensitivity to predictor interpolation and the allocation of Llama-reclassified encounters (Supplementary Sections 7, 11 and 13).

### Performance Evaluation: Feature Importance

To assess feature importance in our model, we used SHAP values, which quantify how much each input feature contributes to the model’s output [28]. Because our inputs consist of sequential clinical data aggregated into 8-hour intervals, SHAP values were computed for each 8-hour interval within the input window. For the global ranking, absolute SHAP values were averaged across the intervals in the window and then across observation windows, and features were ordered by this mean absolute value. For display, each observation window contributes one point per feature, obtained by averaging the signed SHAP values across its intervals so that the direction of effect is preserved. The top 15 features were visualized using a custom SHAP plot, which illustrates both the strength and direction of each feature’s influence on the model’s output, along with the normalized input values. This analysis offers interpretable insights into the model’s decision-making process and highlights the most influential features driving the detection of sepsis.

### Ethics declarations

This retrospective observational study was approved by the Institutional Review Board of Hartford HealthCare. An IRB-approved study protocol was in place. The requirement for informed consent was waived due to the retrospective nature of the study and the use of existing patient-level clinical data. The study was not registered. Patients and members of the public were not involved in the study. The study was conducted in accordance with the Declaration of Helsinki.

## Data availability

The data used in this study were obtained from seven hospitals within the Hartford HealthCare system in the United States and are subject to the Health Insurance Portability and Accountability Act (HIPAA). Because the dataset contains protected health information and other sensitive patient-level clinical information, and because of the data use agreement with Hartford HealthCare, the data cannot be made publicly available. Access to the data requires appropriate regulatory approvals, including HIPAA training or certification and authorization by the relevant institutional review boards and collaborating healthcare system.

## Code availability

The code developed for this study is publicly available on GitHub at https://github.com/ ClaudioAmezaga/sepsis-ml-prediction. The underlying clinical data are not publicly available due to privacy and institutional access restrictions. Model development and testing were conducted in Python 3.12 using publicly available packages, including pandas, numpy, scikit-learn (version 1.7.2), xgboost (version 3.0.5), transformers, torch, datasets, evaluate, matplotlib, shap, and fire.

## Data Availability

Access to the data requires appropriate regulatory approvals, including HIPAA training or certification and authorization by the relevant institutional review boards and collaborating healthcare system.

## Acknowledgements

Hartford HealthCare provided access to the clinical data used in the study. Hartford HealthCare investigators contributed to clinical case reassessment, interpretation of the analyses, and review of the manuscript.

## Author Contributions

I.K.K. co-led model development and evaluation, co-led the data analysis, and drafted and edited the manuscript. C.P.A. co-led model development and evaluation, co-led the data analysis, and drafted and edited the manuscript. E.S.L. co-led study design and scope; provided clinical insight; and participated in manuscript writing and review. A.P. co-led study design and scope; provided clinical insight; and participated in manuscript writing and review. J.G. co-led study design and scope; provided clinical insight; and participated in manuscript writing and review. U.S.W. co-led study design and scope; provided clinical insight; and participated in manuscript writing and review. A.O. co-led the study design and coordination, guided the data preprocessing and the machine learning model development, actively participated in and led the manuscript writing and revision process. All authors read and approved the final manuscript.

