## Supplementary Information for "Language model–assisted label refinement for accurate sepsis detection from electronic health records"

### 1 Epic Score

All results presented in this manuscript that involve the Epic score are based exclusively on the subset of encounters for which the Epic score [1] was available in our database. We did not compute the Epic score ourselves; rather, it was extracted as recorded in the electronic health record. The characteristics of this population are detailed in Supplementary Table S3. A recorded score was available for 330,476 of 352,006 derivation encounters (93.9%) and for 324 of 349 physician-adjudicated encounters. Among derivation encounters, the corresponding numbers were 53,399 of 57,301 septic, 142,056 of 150,807 non-septic SIRS+ and 135,021 of 143,898 non-septic SIRS− encounters.

To ensure consistency in temporal resolution, we aggregated the Epic score into 8-hour intervals, calculating the mean score within each interval. This approach matches the aggregation method used for our own features, allowing for direct comparison across analyses. For evaluation, Epic scores were min–max normalized using the full range of recorded scores (0–93.13). Operating thresholds are presented on the normalized scale, together with the corresponding recorded values. The precise version of the Epic model deployed during the study period was not available.

### 2 Diagnosis Codes

To systematically identify sepsis cases within our EHR dataset, we used International Classification of Diseases, Tenth Revision, Clinical Modification (ICD-10-CM) diagnostic codes associated with sepsis-related diagnoses. An encounter was considered sepsis-coded if a diagnosis recorded during the encounter, or its primary coded diagnosis, was an EHR diagnosis named as sepsis. Each such diagnosis maps to one or more ICD-10-CM codes; all 41 codes are listed in Supplementary Table S16. In the final septic derivation cohort, A41.9 (sepsis, unspecified organism) accounted for 90.8% of encounters, and each organism-specific code for fewer than 10%. The codes identified potentially septic encounters; the SIRS and infection-investigation criteria were then used to determine their positive 8-hour intervals.

### 3 Systemic Inflammatory Response Syndrome (SIRS)

Systemic Inflammatory Response Syndrome (SIRS) is a clinical syndrome characterized by a generalized inflammatory state affecting the whole body, which may be triggered by infectious or noninfectious insults such as trauma, surgery, or pancreatitis. SIRS is defined by the presence of at least two of four clinical criteria [2]. We evaluated SIRS using the following measurements:

- Body temperature >38°C (100.4°F) or <36°C (96.8°F)

- Heart rate >90 beats per minute
- Respiratory rate >20 breaths per minute
- White blood cell count $>12 \times 10^9$/L or $<4 \times 10^9$/L

These criteria were established to provide a sensitive screening tool for identifying patients with systemic inflammation, particularly in the context of suspected infection. However, SIRS can also occur in response to noninfectious causes. In our implementation, the four components were assessed from measurements recorded or interpolated within each 8-hour interval, before multivariable imputation. A missing component was counted as not meeting its criterion, and at least two positive components were required. The same rule was used to establish the SIRS component of the operational labels and to evaluate SIRS as a comparator.

### 4 SOFA Implementation

The SOFA comparator was calculated for each 8-hour interval from the interval means of the required measurements, using the organ-system thresholds of the original score [6] with the adaptations described here. Because SOFA is conventionally calculated using the worst values within a period, our use of interval means may underestimate the severity of organ dysfunction. Respiratory points (1–4) were assigned for ratios of arterial $PaO_2$ (mmHg) to $FiO_2$ below 400, 300, 200 and 100; ventilation status was not extracted, so the requirement for respiratory support at 3 and 4 points was not applied. Coagulation points were assigned for platelet counts below 150, 100, 50 and 20 $\times 10^9$/L, and liver points for total bilirubin of 1.2–1.9, 2.0–5.9, 6.0–12.0 and above 12.0 mg/dL.

Cardiovascular scoring assigned 1 point for a non-invasive mean arterial pressure below 70 mmHg; in these intervals, dopamine ≤5 or any dobutamine gave 2 points, dopamine >5 or adrenaline or noradrenaline ≤0.1 gave 3 points, and dopamine >15 or adrenaline or noradrenaline >0.1 $\mu$g/kg/min gave 4 points. Vasopressor doses were taken from medication administration records recorded in $\mu$g/min or $\mu$g/kg/min, converted to $\mu$g/kg/min using body weight and averaged within each interval; intervals without a recorded administration were treated as receiving none, and vasopressor criteria were applied only when mean arterial pressure was below 70 mmHg.

Central nervous system points were assigned for Glasgow Coma Scale scores of 13–14, 10–12, 6–9 and below 6. Renal points were assigned for creatinine of 1.2–1.9, 2.0–3.4, 3.5–4.9 and at least 5.0 mg/dL, or for urine output below 500 or 200 mL/day (3 and 4 points), with daily output estimated as three times the total recorded in the interval.

Glasgow Coma Scale, urine output, $FiO_2$, mean arterial pressure and weight were obtained from flowsheet documentation, and $PaO_2$, platelets, bilirubin and creatinine from laboratory results (Supplementary Table S14). Inputs that are also STRIDE predictors used the training-fitted imputation described in Methods; $PaO_2$, $FiO_2$, Glasgow Coma Scale, urine output, mean arterial pressure, weight and lactate were completed by a separate imputer fitted on training patients only, with values constrained to be non-negative. Lactate was imputed as an auxiliary variable and not scored. For discrimination and calibration analyses, the integer score was min–max normalized; operating thresholds are reported in points.

### 5 Infection-investigation Criterion and Clinical Variable Sources

Culture orders comprised 107 microbiology culture components in the laboratory results and were assigned to 8-hour intervals by their order date and time. Imaging included any study recorded in the EHR imaging table, regardless of modality or body region, and was assigned to all three intervals of its order date. A positive label can therefore precede the imaging order by up to 16 hours.

In the derivation cohort, the infection-investigation criterion for the 404,671 positive intervals was met by imaging alone in 75.9%, culture alone in 3.5% and both in 20.5%. The corresponding proportions in the physician-adjudicated cohort were 77.8%, 3.9% and 18.3%. Among positive intervals with imaging, radiographs were present in 71.4%, computed tomography in 31.5% and chest studies in 69.7%; the most frequent was a portable chest radiograph (57.4%).

The imaging table also contained electrocardiograms, electroencephalograms, echocardiograms and vascular duplex examinations. Studies of these types, without a culture or radiological study on the same day, accounted for 4.7% of positive derivation intervals (0.5% supported only by electrocardiograms or electroencephalograms) and 4.1% of positive adjudicated intervals. Such studies were the only basis for positive intervals in 186 derivation encounters and no adjudicated encounters.

### 6 Missing Data Imputation

Following the initial linear interpolation step, missing data rates were generally low for the routinely collected vital signs (Supplementary Table S2). In the derivation cohort, these ranged from 3.8% to 6.3% across the three encounter groups, whereas some laboratory measurements were missing more frequently, particularly among non-septic encounters. To address these remaining missing values, `IterativeImputer` [3] was applied using a multivariate imputation strategy inspired by the Multivariate Imputation by Chained Equations (MICE) method [4].

In this approach, each feature with missing values is treated as a regression problem, with the other features used as predictors. Initially, missing values are filled using a simple imputation strategy. The regression models are then fitted sequentially, using the latest imputed values of the other variables, and the process is repeated over several iterations. For each patient-level split, we fitted one imputer to pooled training observations from both outcome classes, sampling at most 200,000 training rows and performing ten iterations. The fitted imputer was subsequently applied to the evaluation cohorts without refitting. The additional SOFA variables were processed with a separate training-fitted imputer.

Interval labels were constructed from recorded and interpolated measurements before multivariable imputation. Linear interpolation within an encounter could use measurements obtained later in the hospital stay. The multivariable imputer was fitted using training patients and then applied to the test and adjudicated cohorts. We examined the effect of omitting predictor interpolation below. The characteristics of included and excluded encounters are compared in Supplementary Table S15.

### 7 Sensitivity Analysis without Linear Interpolation

A predictor-level sensitivity analysis omitted linear interpolation and used the training-fitted multivariable imputer to complete missing predictor values. Encounters, patient partitions, 8-hour intervals and outcome labels were identical to the main analysis; the labels remained derived from recorded and interpolated measurements.

When SIRS was evaluated on recorded values only, 81.9% of positive intervals in the derivation cohort and 80.9% in the physician-adjudicated cohort still met at least two SIRS criteria, and 98.7% and 99.3% of positive encounters retained at least one such interval; three or four of the four SIRS components were recorded in 95.0% and 93.7% of positive intervals, respectively.

Without predictor interpolation, STRIDE ROC-AUC decreased from 0.949 to 0.922 (8 hours), 0.949 to 0.939 (24 hours) and 0.945 to 0.935 (48 hours) in derivation testing, and from 0.871 to 0.844, 0.870 to 0.856 and 0.861 to 0.848 in the physician-adjudicated cohort (Supplementary Table S10). SIRS and Epic results were unchanged because the sensitivity analysis removed interpolation only from STRIDE predictors. STRIDE retained the highest ROC-AUC in derivation testing for all three windows; in the physician-adjudicated cohort, STRIDE-8 (0.844) was slightly below SIRS (0.848), with smaller reductions for the 24- and 48-hour models than for STRIDE-8.

In this analysis, some imputed body weights were truncated at zero by the non-negativity constraint of the SOFA-specific imputer, so weight-based vasopressor doses could not be calculated for the affected intervals; this limits interpretation of the SOFA comparator in this analysis only. No recorded weight was zero, and in the main analysis no interval with a truncated weight had a vasopressor dose requiring weight-based conversion.

### 8 Llama Application

To enhance the reliability of sepsis labeling in our cohort, we employed the Llama 3.1-8B Instruct language model [5] for automated review of discharge summaries. Before prompt development began, infectious disease specialists had completed adjudication of 349 randomly selected, initially sepsis-positive encounters. They confirmed sepsis in 306 and rejected it in 43. Physicians reviewed the complete clinical record, including diagnosis codes, and one final label was retained per encounter. Independent paired labels were not available for calculating inter-rater agreement.

Llama 3.1-8B Instruct was run locally on Hartford HealthCare GPU servers using the Hugging Face `transformers` library in half precision, so no clinical text left the health-system environment. Each encounter's discharge summary was processed individually, with output limited to three tokens and the model's default generation settings (temperature 0.6, top-$p$ 0.9); the first "Yes" or "No" in the response was taken as the label. The model received the *Hospital Course* section, identified from section headings; when no such heading was found, the hospital-stay, history-of-present-illness or discharge-diagnosis section was used, and the complete summary only when none of these could be identified. Notes were not truncated. We developed the prompt using the completed physician labels and evaluated agreement on these same encounters (Supplementary Table S12). The final prompt was then applied to initially positive derivation encounters. Initially non-septic encounters were not reassessed.

The prompt instructed the model to identify explicit evidence of infection or sepsis as a primary issue during the hospital stay or at admission, relying exclusively on information present in the provided clinical notes. The system prompt included detailed inclusion and exclusion criteria, as shown below:

```
{
  "role": "system",
  "content": (
    "You are a medical assistant trained to analyze clinical notes for evidence of
       infection or sepsis. Your task is to determine if the patient currently has
       or previously had a confirmed infection or sepsis based solely on the
       provided notes.\n\n"
    "IMPORTANT INSTRUCTION: Analyze the text carefully for ANY indication that the
       patient has or had an infection or sepsis only as primary issue during the
       hospital stay or at the admission. Be thorough and consider explicit
       statements that definitively indicates infection.\n\n"
    "Criteria for \"Yes\":\n"
    "1. Explicit statements of confirmed infection or sepsis:\n"
    " - Direct mentions like 'patient has infection', 'diagnosed with sepsis',
```

```
    'positive for [pathogen]', 'culture grew [organism]'\n"
"  - Clear documentation of any infectious disease diagnosis\n"
"  - Phrases like 'treated for infection', 'recovering from sepsis'\n\n"
"2. Laboratory or diagnostic evidence confirming infection:\n"
"   - Positive culture results (blood, urine, wound, sputum, etc.)\n"
"   - Positive tests for specific pathogens (PCR, antigen tests, etc.)\n"
"   - Elevated inflammatory markers WITH clear attribution to
    infection/sepsis\n\n"
"3. Clinical documentation that definitively indicates infection:\n"
"  - Any mention of starting or continuing antibiotics FOR a specific infection
    (not prophylactic)\n"
"   - Documentation of purulent discharge, abscess, or infected wounds\n"
"   - Any mention of treatment response to antimicrobial therapy\n\n"
"4. Common infections (including but not limited to):\n"
"   - Urinary: UTI, pyelonephritis, cystitis\n"
"   - Skin/soft tissue: cellulitis, abscess, wound infection\n"
"   - Abdominal: cholecystitis, cholangitis, diverticulitis, peritonitis, C.
    diff\n"
"   - Bloodstream: bacteremia, sepsis, septic shock, fungemia\n"
"   - CNS: meningitis, encephalitis\n"
"   - Other: osteomyelitis, endocarditis, any infection mentioned by name\n\n"
"Criteria for \"No\":\n"
"1. Suspected but unconfirmed infections:\n"
"   - Phrases like 'rule out infection', 'possible infection', 'suspected
    sepsis'\n"
"   - Differential diagnoses that include infection but don't confirm it\n"
"   - Prophylactic antibiotics without stated infection\n"
"   - Colonized by [possible bacteria] but with no toxin\n\n"
"2. Conditions:\n"
" - 'epilepticus', 'pancreatitis', Ulcerative colitis flare', 'Ischemic
    colitis', 'alcoholic cirrhosis/alcoholic hepatitis', 'anemia', 'Syncope',
    'pleural effusion'\n"
" - 'thrombocytosis', 'hypokalemia','Food aspiration pneumonitis', 'Acute liver
    failure', 'Drug overdose', 'Cardiac arrest', 'Necrotizing pancreatitis'\n"
"  - 'PE', Acute gouty arthritis', 'bowel obstruction', 'stroke', 'aspiration
    pneumonia', 'leukocytosis', 'thalamic intracerebral hemorrhage'\n\n"
"3. Explicitly ruled out infections:\n"
"   - If the phrase 'sepsis was ruled' out exists then answer 'No'\n"
"   - 'No evidence of infection', 'sepsis unlikely', 'cultures negative'\n"
"   - Clear alternative non-infectious diagnosis provided\n\n"
```

```
    "Answer Format:\n"
    "Your entire response must be exactly one word: either Yes or No (without
       quotes).\n"
    "Do not include explanations, reasoning, or additional text."
  )
},
{
  "role": "user",
  "content": (
    "Review the following clinical notes: '{text}'. Based strictly on the criteria
       provided and the information within these notes, does the patient currently
       have or previously had an infection or sepsis? "
    "Reply with exactly one word: Yes or No."
  )
}
```

Llama reviewed encounters retrospectively, while the timing of positive 8-hour intervals was assigned using the operational SIRS and infection-investigation criteria. In the physician-adjudicated cohort, confirmed encounters retained their operational interval labels and all intervals in rejected encounters were negative.

### 9 Sliding-Window Construction

Analyzing time series data often requires methods that can effectively capture and utilize temporal dependencies. The sliding window technique is a widely used strategy for this purpose. By partitioning sequential data into overlapping segments of fixed length, the sliding window approach enables the extraction of localized patterns and trends within the data. The following formal framework outlines how sliding windows are constructed in this study.

Formally, consider a time series of data $\{(X_t, y_t) \mid t = 1, 2, \ldots, T\}$, where:

- $X_t$ is the feature vector at time t, with dimension d,
- $y_t$ is the target variable at time t (indicating whether the patient has sepsis).

For a window of size $n$, the sliding window $W_t$ at time $t$ is defined as:

$$W_t = [X_{t-n+1}, X_{t-n+2}, \ldots, X_t]$$

Each window $W_t$ thus has a dimension of $n \times d$, which can be flattened into a vector of length $nd$ to be used in standard supervised learning models. The transformed dataset is constructed by iterating this process for each t from n to T, yielding:

$$\{(W_n, y_n), (W_{n+1}, y_{n+1}), \ldots, (W_T, y_T)\}$$

This formalization enables the encoding of temporal dependencies in a format compatible with machine learning algorithms.

Here, one time step corresponds to an 8-hour interval. We used $n = 1$, 3 and 6 for the STRIDE-8, STRIDE-24 and STRIDE-48 models. Each window ends at the interval whose sepsis status is being classified, and all three models used the same encounter eligibility requirement of at least 48 hours of recorded data.

### 10 Complete Metrics and Results

In addition to the 8-hour window results presented in the main manuscript, we report complete performance metrics for the 24- and 48-hour windows across all cohorts in Supplementary Table S1 (Supplementary Figs. S1–S3). STRIDE achieved ROC-AUCs of 0.949 and 0.945 in derivation testing for the 24- and 48-hour windows, respectively, compared with 0.870 and 0.861 in the physician-adjudicated cohort. Within septic encounters, the corresponding ROC-AUCs were 0.906 and 0.899. The models retained higher discrimination than SOFA and Epic across these comparisons.

Beyond discrimination, we report precision–recall and calibration metrics for all three windows in Supplementary Table S4: average precision, Brier score and expected calibration error (ECE). In derivation testing, STRIDE average precision was 0.458, 0.477 and 0.423 for the 8-, 24- and 48-hour windows, respectively. Brier scores were 0.040, 0.040 and 0.041. In the physician-adjudicated cohort, average precision was 0.557, 0.566 and 0.512. ROC-AUC and average precision were higher for STRIDE than for the two score-based comparators in all three windows.

The 95% confidence intervals were calculated from 200 patient-level bootstrap resamples within each of five patient-level splits, pooling the 1,000 resampled estimates. Each derivation test set was divided into two patient groups: thresholds were selected on one group and evaluated on the other, with the groups then exchanged. The physician-adjudicated cohort was evaluated using thresholds selected from derivation data. SIRS was assessed at its fixed rule of at least two criteria.

### 11 SIRS Ablation and Subgroup Analyses

To assess the contribution of SIRS-related predictors, STRIDE was retrained after removing the minimum, mean and maximum of temperature, heart rate, respiratory rate and white blood cell count from each 8-hour interval. This removed 12, 36 and 72 input columns from the 8-,

24- and 48-hour models, respectively. The patient partitions, outcome labels, imputed values, scaling and hyperparameters remained unchanged.

With these predictors excluded, ROC-AUC declined from 0.949 to 0.853 in derivation testing and from 0.871 to 0.743 in physician-adjudicated evaluation for STRIDE-8. The corresponding values were 0.949 to 0.868 and 0.870 to 0.751 for STRIDE-24, and 0.945 to 0.867 and 0.861 to 0.745 for STRIDE-48. For comparison, Epic and SOFA achieved ROC-AUCs of 0.828 and 0.742 in derivation testing and 0.724 and 0.637 in the adjudicated cohort.

We also assessed discrimination among intervals already meeting at least two SIRS criteria. This subset contained, on average, 113,081 positive and 399,673 negative intervals per derivation test split and 2,823 positive and 2,884 negative intervals in the adjudicated cohort. STRIDE-8 achieved ROC-AUCs of 0.765 and 0.576, compared with 0.685 and 0.568 for Epic and 0.632 and 0.513 for SOFA, respectively. Average precision was 0.458, 0.390 and 0.299 for STRIDE-8, Epic and SOFA in derivation testing, and 0.557, 0.536 and 0.488 in the physician-adjudicated cohort. STRIDE-8 average precision was unchanged from the full cohorts because nearly every interval not meeting SIRS was ranked below every positive interval; excluding these intervals therefore lowered ROC-AUC without changing precision at any positive interval. Positive intervals represented 22.1% and 49.5% of the two subsets. With SIRS fixed, the remaining variation in the operational label reflected the culture or imaging-order criterion within sepsis-coded encounters. Epic results were calculated for intervals with a recorded score.

STRIDE-8 discrimination was similar across hospitals, calendar years and demographic groups (Supplementary Table S11). In the derivation test set, ROC-AUC ranged from 0.938 to 0.967 across the seven hospitals, from 0.943 to 0.952 across calendar years 2019–2025, and from 0.945 to 0.954 across sex, age-band, race and ethnicity groups with recorded values. Average precision varied more (0.407–0.544 across hospitals; 0.505 in 2019 and 0.436 in 2025), in part reflecting differences in the proportion of positive intervals (4.2–7.1% across hospitals; 6.4% and 4.6% in 2019 and 2025).

### 12 Random Forest vs XGBoost

Table S5 presents a comparison of ROC-AUC scores between XGBoost and Random Forest models for sepsis detection using 8-, 24- and 48-hour observation windows across multiple cohorts. Random Forest used 100 trees and a maximum depth of 12, with the same patient partitions and preprocessed features as STRIDE. The two methods performed similarly with an 8-hour observation window: Random Forest achieved ROC-AUCs of 0.951 and 0.870 in derivation testing and physician-adjudicated evaluation, compared with 0.949 and 0.871 for STRIDE-8. XGBoost performed slightly better with the 24- and 48-hour windows, particularly in the adjudicated cohort (Supplementary Table S5).

### 13 Sensitivity to the Allocation of Llama-Reclassified Encounters

In the main analysis, patients with a Llama-reclassified encounter were assigned to training. We examined two alternative designs using five patient-level 70/30 splits of the 224,756 derivation patients, with the same imputation, scaling and hyperparameters. In design A, these patients were allocated in the same way as other patients, retaining the negative labels assigned by Llama. In design B, reclassified encounters were excluded from both training and testing. Design A placed an average of 2,551 reclassified encounters in the test set per split.

ROC-AUC in the physician-adjudicated cohort was 0.872 for both designs, compared with 0.871 in the main analysis (Supplementary Table S13). In design A, derivation-test ROC-AUC was 0.943 with the refined labels, 0.948 after excluding reclassified encounters from evaluation and 0.946 when those encounters were evaluated under their original code-based labels. At the threshold targeting 80% sensitivity in the remaining test encounters, STRIDE-8 classified as positive 74.5% of intervals in the reclassified encounters that met the operational criteria (18,719 intervals per split, on average).

### 14 Additional Operating Points

At thresholds selected to target 80% sensitivity in derivation testing, STRIDE-24 achieved a specificity of 91.1% and an accuracy of 90.4% in derivation testing (Supplementary Table S6). In the physician-adjudicated cohort, specificity and accuracy were 77.9% and 78.5%. For STRIDE-48, the corresponding values were 90.5% and 89.9% in derivation testing and 76.6% and 77.5% in the adjudicated cohort (Supplementary Table S7). SIRS was evaluated at its fixed clinical rule, whereas the other operating points were selected from derivation data.

To further characterize model performance at different operating points, we report sensitivity targets of 85% and 90% for all three observation windows. Tables S8 and S9 summarize the thresholds, specificity and accuracy in derivation testing and the physician-adjudicated cohort. Higher sensitivity targets required lower thresholds and generally reduced specificity. The thresholds reported for Epic reflect the min–max normalized recorded scores, with their corresponding original values provided for reference.

### Supplementary Figures

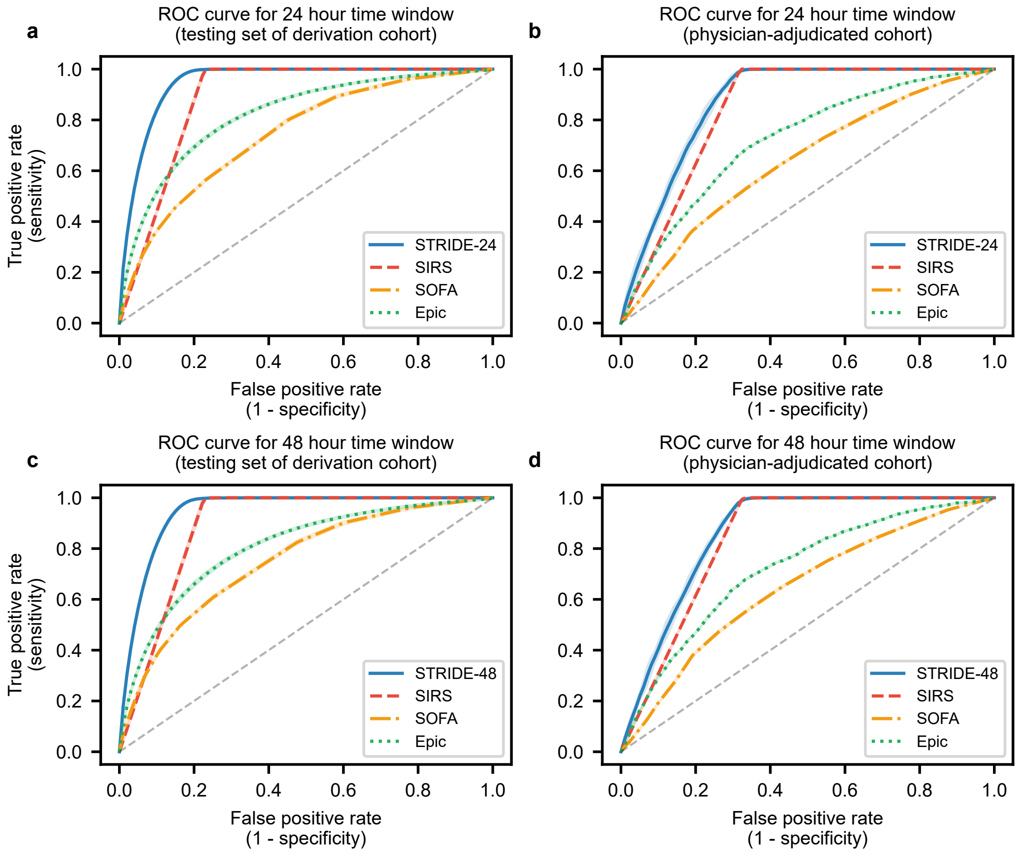


Supplementary Fig. S1: **Receiver operating characteristic curves for the 24- and 48-hour windows.** ROC curves for STRIDE and the clinical benchmarks (SOFA, Epic, and SIRS) for the 24-hour (**a**, **b**) and 48-hour (**c**, **d**) windows, on the testing set of the derivation cohort (**a**, **c**) and the physician-adjudicated cohort (**b**, **d**). Solid lines show the mean across the five patient-level splits and shaded bands the spread across splits; the dashed diagonal denotes chance.

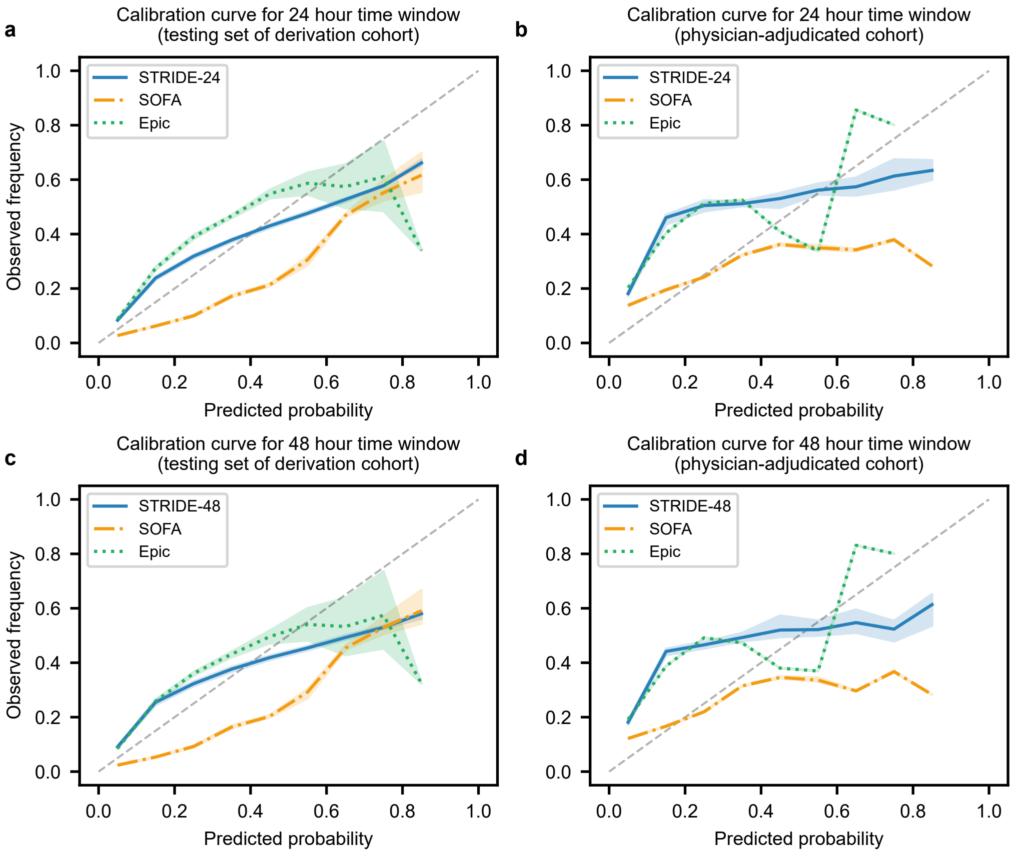


Supplementary Fig. S2: **Calibration curves for the 24- and 48-hour windows.** Calibration curves for STRIDE and the clinical benchmarks (SOFA and Epic) for the 24-hour (**a**, **b**) and 48-hour (**c**, **d**) windows, on the testing set of the derivation cohort (**a**, **c**) and the physician-adjudicated cohort (**b**, **d**). Solid lines show the mean across the five patient-level splits and shaded bands the spread across splits; the dashed diagonal denotes perfect calibration. SOFA and Epic are shown on min–max normalized scores; SIRS is omitted because it is a binary rule. The horizontal axis denotes predicted probability for STRIDE and normalized scores for Epic and SOFA.

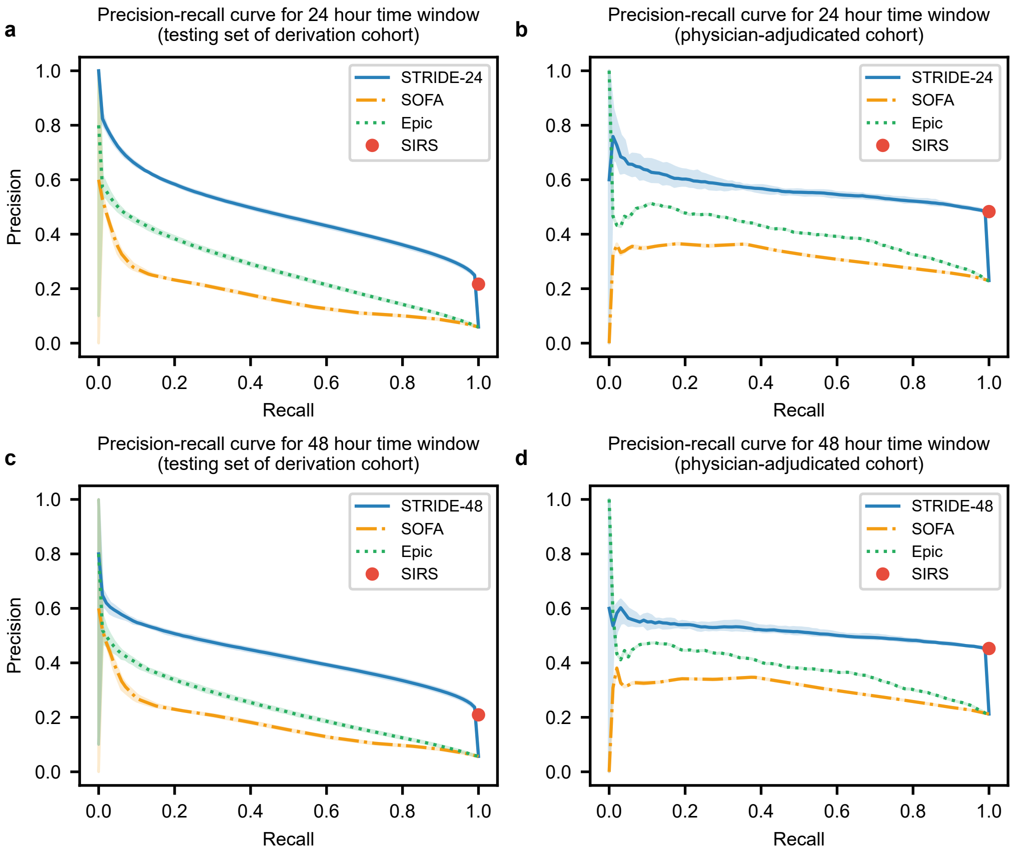


Supplementary Fig. S3: **Precision-recall curves for the 24- and 48-hour windows.** Precision-recall curves for STRIDE and the clinical benchmarks (SOFA, Epic and SIRS) for the 24-hour (**a**, **b**) and 48-hour (**c**, **d**) windows, on the testing set of the derivation cohort (**a**, **c**) and the physician-adjudicated cohort (**b**, **d**). Solid lines show the mean across the five patient-level splits and shaded bands the spread across splits. SIRS is shown as a red circle at its fixed operating point.

### Supplementary Tables

Supplementary Table S1: ROC-AUC with 95% confidence intervals (patient-level bootstrap) for sepsis detection methods using 24-hour and 48-hour time windows. The septic subset contains positive and negative intervals from septic encounters; the mixed groups additionally include intervals from their respective non-septic encounter groups.

| **Time Window** | **Method** | **Testing Set of Derivation Cohort** | **Septic Subset of Derivation Cohort** | **Septic + non-septic SIRS+ encounters** | **Septic + non-septic SIRS− encounters** | **Physician-adjudicated cohort** |
|---|---|---|---|---|---|---|
| 24-Hour | STRIDE-24 | 0.949 [0.948–0.951] | 0.906 [0.902–0.910] | 0.928 [0.926–0.930] | 0.961 [0.959–0.963] | 0.870 [0.830–0.903] |
| | SOFA | 0.744 [0.737–0.750] | 0.669 [0.661–0.677] | 0.709 [0.702–0.716] | 0.762 [0.756–0.768] | 0.635 [0.583–0.681] |
| | Epic | 0.822 [0.815–0.829] | 0.721 [0.713–0.728] | 0.784 [0.776–0.791] | 0.834 [0.827–0.840] | 0.720 [0.670–0.762] |
| | SIRS | 0.886 [0.884–0.888] | 0.870 [0.866–0.874] | 0.844 [0.842–0.847] | 0.937 [0.935–0.939] | 0.840 [0.807–0.870] |
| 48-Hour | STRIDE-48 | 0.945 [0.943–0.947] | 0.899 [0.895–0.903] | 0.926 [0.923–0.928] | 0.953 [0.950–0.955] | 0.861 [0.819–0.896] |
| | SOFA | 0.750 [0.742–0.758] | 0.688 [0.679–0.695] | 0.721 [0.713–0.729] | 0.765 [0.758–0.772] | 0.644 [0.588–0.692] |
| | Epic | 0.805 [0.797–0.813] | 0.709 [0.700–0.717] | 0.771 [0.763–0.780] | 0.811 [0.803–0.818] | 0.717 [0.662–0.764] |
| | SIRS | 0.886 [0.884–0.889] | 0.869 [0.864–0.873] | 0.852 [0.849–0.854] | 0.930 [0.927–0.933] | 0.837 [0.802–0.868] |

Supplementary Table S2: Percentage of 8-hour intervals with a missing value, by population group and clinical variable, after linear interpolation and before multivariable imputation.

| | Derivation Cohort | | | Physician-adjudicated cohort | |
|---|---|---|---|---|---|
| **Variable** | **Septic** | **Non-septic SIRS+** | **Non-septic SIRS−** | **Septic** | **Non-septic SIRS+** |
| Heart rate | 3.8% | 4.7% | 6.0% | 3.5% | 2.3% |
| Respiratory rate | 3.8% | 4.8% | 6.1% | 3.5% | 2.3% |
| Systolic BP | 3.9% | 4.8% | 6.0% | 3.5% | 2.9% |
| Diastolic BP | 3.9% | 4.8% | 6.0% | 3.5% | 2.9% |
| Temperature | 3.9% | 4.8% | 6.1% | 3.5% | 2.4% |
| Oxygen saturation | 3.8% | 4.8% | 6.3% | 3.5% | 2.3% |
| Bilirubin | 7.8% | 18.0% | 22.7% | 7.5% | 5.0% |
| Alanine Aminotransferase | 8.0% | 18.8% | 22.0% | 7.3% | 5.0% |
| Aspartate Aminotransferase | 6.9% | 18.0% | 20.9% | 6.4% | 5.0% |
| Alkaline phosphatase | 6.7% | 17.5% | 20.6% | 6.0% | 5.0% |
| Blood Urea Nitrogen | 4.1% | 5.9% | 7.8% | 3.8% | 2.5% |
| Creatinine | 4.1% | 5.8% | 7.7% | 3.7% | 2.5% |
| Platelets | 4.2% | 5.9% | 8.4% | 3.9% | 2.6% |
| Potassium | 4.2% | 6.1% | 7.9% | 3.7% | 2.5% |
| Sodium | 4.1% | 5.9% | 7.8% | 3.7% | 2.5% |
| Chloride | 4.1% | 5.9% | 7.8% | 3.7% | 2.5% |
| Albumin | 6.4% | 16.9% | 19.7% | 5.2% | 5.0% |
| White Blood Cell Count | 4.1% | 5.8% | 8.3% | 3.8% | 2.6% |
| $CO_2$ | 4.1% | 5.9% | 7.8% | 3.7% | 2.5% |

Supplementary Table S3: Characteristics of the encounters with a recorded Epic score (Epic-eligible population), defined as in Table 1 of the main text.

| | Derivation Cohort | | | Physician-adjudicated cohort | |
|---|---|---|---|---|---|
| **Variable** | **Septic** | **Non-septic SIRS+** | **Non-septic SIRS−** | **Septic** | **Non-septic SIRS+** |
| **Population** | | | | | |
| Number of Patients | 39,610 | 107,699 | 96,526 | 283 | 41 |
| Number of Encounters | 53,399 | 142,056 | 135,021 | 283 | 41 |
| **Demographics** | | | | | |
| Age at admission, mean (SD) | 67.2 (16.8) | 63.1 (19.1) | 63.6 (19.3) | 67.8 (16.6) | 60.6 (18.7) |
| Men, n (%) | 28,048 (52.5%) | 68,725 (48.4%) | 61,847 (45.8%) | 151 (53.4%) | 19 (46.3%) |
| Women, n (%) | 25,136 (47.1%) | 72,820 (51.3%) | 72,589 (53.8%) | 129 (45.6%) | 21 (51.2%) |
| Unknown, n (%) | 215 (0.4%) | 511 (0.4%) | 585 (0.4%) | 3 (1.1%) | 1 (2.4%) |
| **Race, n (%)** | | | | | |
| White or Caucasian | 39,974 (74.9%) | 101,948 (71.8%) | 97,543 (72.2%) | 218 (77.0%) | 28 (68.3%) |

Continued on next page

*Table S3 (continued)*

| | Derivation Cohort | | | Physician-adjudicated cohort | |
|---|---|---|---|---|---|
| **Variable** | **Septic** | **Non-septic SIRS+** | **Non-septic SIRS−** | **Septic** | **Non-septic SIRS+** |
| Other | 7,992 (15.0%) | 22,132 (15.6%) | 20,672 (15.3%) | 28 (9.9%) | 7 (17.1%) |
| Black or African American | 4,726 (8.9%) | 15,682 (11.0%) | 14,931 (11.1%) | 31 (11.0%) | 5 (12.2%) |
| Unknown | 707 (1.3%) | 2,294 (1.6%) | 1,875 (1.4%) | 6 (2.1%) | 1 (2.4%) |
| **Ethnicity, n (%)** | | | | | |
| Not Hispanic or Latino | 45,269 (84.8%) | 119,102 (83.8%) | 113,522 (84.1%) | 249 (88.0%) | 31 (75.6%) |
| Hispanic or Latino | 7,268 (13.6%) | 20,128 (14.2%) | 19,205 (14.2%) | 29 (10.2%) | 7 (17.1%) |
| Unknown | 862 (1.6%) | 2,826 (2.0%) | 2,294 (1.7%) | 5 (1.8%) | 3 (7.3%) |
| **Smoking status, n (%)** | | | | | |
| Never smoker | 19,830 (37.1%) | 54,826 (38.6%) | 55,428 (41.1%) | 110 (38.9%) | 13 (31.7%) |
| Former smoker | 20,061 (37.6%) | 45,742 (32.2%) | 42,839 (31.7%) | 91 (32.2%) | 15 (36.6%) |
| Current smoker | 7,264 (13.6%) | 20,637 (14.5%) | 20,427 (15.1%) | 47 (16.6%) | 6 (14.6%) |
| Never assessed | 1,705 (3.2%) | 5,825 (4.1%) | 4,405 (3.3%) | 10 (3.5%) | 2 (4.9%) |
| Other / unknown | 4,539 (8.5%) | 15,026 (10.6%) | 11,922 (8.8%) | 25 (8.8%) | 5 (12.2%) |
| **Admission type, n (%)** | | | | | |
| Emergency | 49,336 (92.4%) | 107,558 (75.7%) | 108,313 (80.2%) | 251 (88.7%) | 31 (75.6%) |
| Urgent | 2,730 (5.1%) | 8,639 (6.1%) | 8,806 (6.5%) | 18 (6.4%) | 8 (19.5%) |
| Elective | 765 (1.4%) | 19,849 (14.0%) | 15,704 (11.6%) | 8 (2.8%) | 0 (0.0%) |
| Other | 568 (1.1%) | 6,010 (4.2%) | 2,198 (1.6%) | 6 (2.1%) | 2 (4.9%) |
| **Length of stay (days), n (%)** | | | | | |
| 0–2 | 5,116 (9.6%) | 25,808 (18.2%) | 43,176 (32.0%) | 21 (7.4%) | 3 (7.3%) |
| 3–5 | 19,674 (36.8%) | 59,777 (42.1%) | 59,760 (44.3%) | 84 (29.7%) | 8 (19.5%) |
| 6–8 | 11,402 (21.4%) | 26,480 (18.6%) | 17,473 (12.9%) | 75 (26.5%) | 14 (34.1%) |
| 9+ | 17,207 (32.2%) | 29,991 (21.1%) | 14,612 (10.8%) | 103 (36.4%) | 16 (39.0%) |

Continued on next page

*Table S3 (continued)*

| | Derivation Cohort | | | Physician-adjudicated cohort | |
|---|---|---|---|---|---|
| **Variable** | **Septic** | **Non-septic SIRS+** | **Non-septic SIRS−** | **Septic** | **Non-septic SIRS+** |
| **Vital Signs, mean (SD)** | | | | | |
| Heart rate (bpm) | 85.5 (11.8) | 82.8 (11.4) | 75.1 (10.5) | 85.4 (11.8) | 89.6 (14.5) |
| Respiratory rate (breaths/min) | 19.2 (2.3) | 18.6 (1.8) | 17.8 (1.0) | 19.5 (2.6) | 19.7 (2.4) |
| Systolic BP (mmHg) | 125.3 (14.6) | 128.0 (14.9) | 131.1 (15.8) | 123.3 (14.8) | 123.3 (14.0) |
| Diastolic BP (mmHg) | 69.0 (7.7) | 71.1 (8.3) | 72.0 (8.3) | 68.0 (8.1) | 68.7 (8.8) |
| Temperature (°F) | 98.3 (0.7) | 98.0 (0.7) | 97.9 (0.5) | 98.3 (0.8) | 98.3 (0.8) |
| Oxygen saturation (%) | 95.9 (1.7) | 96.2 (1.7) | 96.6 (1.6) | 95.8 (1.6) | 96.2 (1.5) |
| **Lab Results, mean (SD)** | | | | | |
| Bilirubin (mg/dL) | 0.8 (1.8) | 0.8 (1.8) | 0.7 (1.1) | 1.0 (1.7) | 1.2 (2.4) |
| Alanine Aminotransferase (U/L) | 32.7 (29.1) | 33.3 (29.8) | 28.8 (27.0) | 35.3 (29.8) | 38.4 (34.0) |
| Aspartate Aminotransferase (U/L) | 38.6 (30.0) | 39.3 (31.2) | 32.8 (26.7) | 42.7 (31.6) | 48.7 (35.0) |
| Alkaline phosphatase (U/L) | 113.0 (61.8) | 102.9 (55.5) | 95.1 (49.5) | 110.9 (60.9) | 115.4 (62.1) |
| Blood Urea Nitrogen (mg/dL) | 25.6 (19.2) | 21.1 (16.1) | 18.5 (13.5) | 29.0 (23.0) | 21.8 (14.5) |
| Creatinine (mg/dL) | 1.3 (1.3) | 1.2 (1.2) | 1.1 (1.1) | 1.4 (1.3) | 1.2 (0.9) |
| Platelets ($\times 10^9$/L) | 238.9 (114.4) | 235.9 (99.0) | 232.4 (84.9) | 213.4 (109.1) | 242.0 (118.9) |
| Potassium (mmol/L) | 4.0 (0.4) | 4.0 (0.4) | 4.0 (0.4) | 4.0 (0.5) | 4.0 (0.3) |
| Sodium (mmol/L) | 138.5 (3.8) | 138.0 (3.4) | 138.4 (3.2) | 138.7 (4.2) | 138.4 (4.6) |
| Chloride (mmol/L) | 103.2 (5.0) | 102.3 (4.5) | 103.0 (4.1) | 103.4 (5.5) | 103.4 (5.0) |
| Albumin (g/dL) | 3.1 (1.1) | 3.6 (0.6) | 3.8 (0.5) | 3.1 (0.6) | 3.1 (0.6) |
| White Blood Cell Count ($\times 10^9$/L) | 11.5 (6.1) | 10.1 (5.4) | 7.8 (2.9) | 11.6 (5.7) | 11.5 (4.7) |
| $CO_2$ (mmol/L) | 24.6 (3.9) | 24.9 (3.7) | 25.1 (3.2) | 24.4 (3.6) | 23.6 (2.9) |
| **Other, mean (SD)** | | | | | |
| Height (cm) | 168.1 (11.4) | 167.9 (11.1) | 167.7 (11.1) | 167.3 (12.1) | 169.0 (11.2) |
| Weight (kg) | 83.8 (27.0) | 83.0 (24.3) | 82.0 (23.1) | 84.1 (24.6) | 81.8 (26.0) |

Supplementary Table S4: Average precision (AP), Brier score and expected calibration error (ECE) with 95% confidence intervals (patient-level bootstrap) for STRIDE and clinical scores in the testing set of the derivation cohort and the physician-adjudicated cohort. Lower Brier and ECE are preferable for the evaluated scores; SOFA and Epic values were computed after min–max normalization of their recorded scores, not after fitting a probability-calibration model. Their Brier scores and ECEs therefore describe these normalized scores and should not be interpreted as calibrated clinical event probabilities.

| Time window | Method | Testing set of derivation cohort | | | Physician-adjudicated cohort | | |
|---|---|---|---|---|---|---|---|
| | | **AP** | **Brier** | **ECE** | **AP** | **Brier** | **ECE** |
| 8-Hour | STRIDE-8 | 0.458 [0.445–0.470] | 0.040 [0.038–0.041] | 0.015 [0.014–0.016] | 0.557 [0.447–0.655] | 0.143 [0.122–0.163] | 0.105 [0.085–0.128] |
| | SOFA | 0.159 [0.151–0.167] | 0.060 [0.058–0.062] | 0.067 [0.064–0.069] | 0.314 [0.242–0.393] | 0.178 [0.161–0.194] | 0.063 [0.046–0.100] |
| | Epic | 0.277 [0.267–0.287] | 0.052 [0.050–0.054] | 0.024 [0.022–0.027] | 0.416 [0.336–0.492] | 0.194 [0.164–0.228] | 0.166 [0.133–0.206] |
| 24-Hour | STRIDE-24 | 0.477 [0.466–0.488] | 0.040 [0.039–0.042] | 0.017 [0.015–0.018] | 0.566 [0.463–0.658] | 0.144 [0.124–0.165] | 0.106 [0.087–0.132] |
| | SOFA | 0.162 [0.153–0.171] | 0.062 [0.060–0.064] | 0.069 [0.066–0.071] | 0.313 [0.247–0.391] | 0.179 [0.161–0.196] | 0.066 [0.047–0.100] |
| | Epic | 0.266 [0.256–0.277] | 0.051 [0.049–0.053] | 0.022 [0.020–0.025] | 0.404 [0.323–0.485] | 0.189 [0.160–0.220] | 0.159 [0.127–0.195] |
| 48-Hour | STRIDE-48 | 0.423 [0.408–0.438] | 0.041 [0.039–0.043] | 0.020 [0.018–0.022] | 0.512 [0.414–0.614] | 0.143 [0.121–0.166] | 0.107 [0.087–0.133] |
| | SOFA | 0.162 [0.152–0.172] | 0.062 [0.060–0.064] | 0.078 [0.075–0.081] | 0.297 [0.227–0.383] | 0.170 [0.150–0.190] | 0.063 [0.041–0.100] |
| | Epic | 0.234 [0.223–0.246] | 0.050 [0.047–0.053] | 0.021 [0.018–0.024] | 0.378 [0.290–0.463] | 0.176 [0.147–0.209] | 0.145 [0.112–0.183] |

Supplementary Table S5: ROC-AUC with 95% confidence intervals (patient-level bootstrap) of XGBoost (STRIDE) and Random Forest models trained on the same patient partitions, imputed and scaled features and labels, for the 8-, 24- and 48-hour windows.

| Time window | Method | Testing set of derivation cohort | Septic subset of derivation cohort | Septic + non-septic SIRS+ encounters | Septic + non-septic SIRS− encounters | Physician-adjudicated cohort |
|---|---|---|---|---|---|---|
| 8-Hour | XGBoost | 0.949 [0.947–0.950] | 0.907 [0.903–0.911] | 0.926 [0.924–0.928] | 0.963 [0.961–0.965] | 0.871 [0.828–0.906] |
| | Random Forest | 0.951 [0.949–0.952] | 0.908 [0.904–0.912] | 0.929 [0.926–0.931] | 0.964 [0.962–0.966] | 0.870 [0.820–0.906] |
| 24-Hour | XGBoost | 0.949 [0.948–0.951] | 0.906 [0.902–0.910] | 0.928 [0.926–0.930] | 0.961 [0.959–0.963] | 0.870 [0.830–0.903] |
| | Random Forest | 0.947 [0.945–0.949] | 0.901 [0.897–0.904] | 0.925 [0.923–0.928] | 0.959 [0.957–0.961] | 0.859 [0.811–0.896] |
| 48-Hour | XGBoost | 0.945 [0.943–0.947] | 0.899 [0.895–0.903] | 0.926 [0.923–0.928] | 0.953 [0.950–0.955] | 0.861 [0.819–0.896] |
| | Random Forest | 0.941 [0.939–0.944] | 0.888 [0.883–0.892] | 0.921 [0.918–0.924] | 0.948 [0.945–0.950] | 0.843 [0.791–0.888] |

Supplementary Table S6: Operating-point performance for the 24-hour window. STRIDE-24, SOFA and Epic thresholds target 80% sensitivity and were selected on derivation data only; SIRS is evaluated at its fixed rule. Values are means over five patient-level splits with 95% confidence intervals (patient-level bootstrap). PPV, positive predictive value; NPV, negative predictive value; TP, FP, TN and FN, true positives, false positives, true negatives and false negatives (classified intervals, means per split). †Mean of the integer SOFA thresholds selected across splits. Epic thresholds are shown on the min–max normalized scale, with the corresponding recorded Epic score in parentheses.

| Method | Cohort | Threshold | Sensitivity | Specificity | PPV | NPV | Accuracy | TP / FP<br>TN / FN |
|---|---|---|---|---|---|---|---|---|
| STRIDE-24 | Testing set of derivation cohort | 10.4% | 80.0%<br>[79.4%–80.7%] | 91.1%<br>[90.8%–91.3%] | 36.1%<br>[34.9%–37.2%] | 98.6%<br>[98.6%–98.7%] | 90.4%<br>[90.1%–90.6%] | 82,477 / 145,937<br>1,487,663 / 20,611 |
| | Physician-adjudicated cohort | 10.4% | 80.4%<br>[76.4%–83.5%] | 77.9%<br>[71.9%–83.1%] | 52.1%<br>[44.1%–59.5%] | 93.0%<br>[91.1%–94.5%] | 78.5%<br>[74.0%–82.5%] | 2,136 / 1,967<br>6,933 / 521 |
| SOFA | Testing set of derivation cohort | 2.3 points† | 86.5%<br>[84.1%–89.6%] | 45.7%<br>[41.5%–49.1%] | 9.2%<br>[8.4%–9.8%] | 98.2%<br>[97.9%–98.5%] | 48.1%<br>[44.2%–51.2%] | 89,167 / 887,019<br>746,581 / 13,920 |
| | Physician-adjudicated cohort | 2.4 points† | 87.3%<br>[79.1%–93.1%] | 26.5%<br>[17.6%–38.5%] | 26.2%<br>[21.5%–31.7%] | 87.6%<br>[84.5%–90.5%] | 40.4%<br>[33.3%–48.8%] | 2,320 / 6,546<br>2,354 / 337 |
| Epic | Testing set of derivation cohort | 3.2%<br>(2.95) | 80.0%<br>[79.0%–81.0%] | 69.4%<br>[68.8%–70.1%] | 14.2%<br>[13.7%–14.8%] | 98.2%<br>[98.1%–98.3%] | 70.0%<br>[69.5%–70.7%] | 77,259 / 465,450<br>1,054,557 / 19,317 |
| | Physician-adjudicated cohort | 3.2%<br>(2.95) | 82.8%<br>[77.4%–87.0%] | 47.0%<br>[40.3%–54.3%] | 31.7%<br>[26.5%–37.0%] | 90.2%<br>[86.9%–92.8%] | 55.2%<br>[49.9%–60.6%] | 2,002 / 4,321<br>3,839 / 416 |
| SIRS | Testing set of derivation cohort | ≥2 criteria | 100.0%<br>[100.0%–100.0%] | 77.2%<br>[76.8%–77.5%] | 21.7%<br>[20.8%–22.5%] | 100.0%<br>[100.0%–100.0%] | 78.5%<br>[78.2%–78.9%] | 103,088 / 372,783<br>1,260,817 / 0 |
| | Physician-adjudicated cohort | ≥2 criteria | 100.0%<br>[100.0%–100.0%] | 68.0%<br>[61.4%–74.0%] | 48.3%<br>[41.5%–54.5%] | 100.0%<br>[100.0%–100.0%] | 75.4%<br>[70.2%–79.7%] | 2,657 / 2,844<br>6,056 / 0 |

Supplementary Table S7: Operating-point performance for the 48-hour window. STRIDE-48, SOFA and Epic thresholds target 80% sensitivity and were selected on derivation data only; SIRS is evaluated at its fixed rule. Values are means over five patient-level splits with 95% confidence intervals (patient-level bootstrap). PPV, positive predictive value; NPV, negative predictive value; TP, FP, TN and FN, true positives, false positives, true negatives and false negatives (classified intervals, means per split). Epic thresholds are shown on the min–max normalized scale, with the corresponding recorded Epic score in parentheses.

| Method | Cohort | Threshold | Sensitivity | Specificity | PPV | NPV | Accuracy | TP / FP<br>TN / FN |
|---|---|---|---|---|---|---|---|---|
| STRIDE-48 | Testing set of derivation cohort | 8.0% | 80.0%<br>[79.2%–80.7%] | 90.5%<br>[90.2%–90.8%] | 33.5%<br>[32.1%–34.8%] | 98.7%<br>[98.6%–98.8%] | 89.9%<br>[89.6%–90.1%] | 63,866 / 126,892<br>1,202,925 / 15,973 |
| | Physician-adjudicated cohort | 8.0% | 80.8%<br>[75.6%–85.1%] | 76.6%<br>[70.2%–82.1%] | 48.1%<br>[40.1%–56.0%] | 93.7%<br>[91.6%–95.3%] | 77.5%<br>[72.3%–81.8%] | 1,785 / 1,924<br>6,283 / 425 |
| SOFA | Testing set of derivation cohort | 3 points | 82.6%<br>[81.6%–83.6%] | 52.3%<br>[51.4%–53.0%] | 9.4%<br>[8.9%–9.9%] | 98.0%<br>[97.9%–98.2%] | 54.0%<br>[53.2%–54.7%] | 65,943 / 634,851<br>694,966 / 13,896 |
| | Physician-adjudicated cohort | 3 points | 84.2%<br>[78.5%–88.7%] | 31.7%<br>[25.3%–38.3%] | 24.9%<br>[20.1%–30.4%] | 88.2%<br>[85.5%–90.6%] | 42.8%<br>[37.6%–48.3%] | 1,860 / 5,604<br>2,603 / 350 |
| Epic | Testing set of derivation cohort | 2.9%<br>(2.65) | 80.0%<br>[78.8%–81.2%] | 65.9%<br>[65.3%–66.7%] | 12.5%<br>[11.9%–13.1%] | 98.2%<br>[98.0%–98.4%] | 66.7%<br>[66.2%–67.5%] | 59,953 / 420,838<br>814,647 / 15,007 |
| | Physician-adjudicated cohort | 2.9%<br>(2.65) | 84.1%<br>[78.3%–88.7%] | 45.0%<br>[37.7%–51.7%] | 29.0%<br>[23.7%–34.1%] | 91.4%<br>[88.0%–94.1%] | 53.2%<br>[47.6%–58.4%] | 1,687 / 4,134<br>3,375 / 319 |
| SIRS | Testing set of derivation cohort | ≥2 criteria | 100.0%<br>[100.0%–100.0%] | 77.3%<br>[76.9%–77.8%] | 20.9%<br>[20.0%–21.8%] | 100.0%<br>[100.0%–100.0%] | 78.6%<br>[78.2%–79.0%] | 79,839 / 301,892<br>1,027,925 / 0 |
| | Physician-adjudicated cohort | ≥2 criteria | 100.0%<br>[100.0%–100.0%] | 67.4%<br>[60.4%–73.6%] | 45.3%<br>[38.1%–52.0%] | 100.0%<br>[100.0%–100.0%] | 74.3%<br>[68.7%–79.1%] | 2,210 / 2,672<br>5,535 / 0 |

Supplementary Table S8: STRIDE performance at 85% target sensitivity across observation windows. Thresholds selected on held-out derivation data as in the main-text operating-point table; means over five patient-level splits with 95% confidence intervals from a patient-level bootstrap. PPV, positive predictive value; NPV, negative predictive value; TP, FP, TN and FN, true positives, false positives, true negatives and false negatives (classified intervals, means per split).

| Method | Cohort | Threshold | Sensitivity | Specificity | PPV | NPV | Accuracy | TP / FP<br>TN / FN |
|---|---|---|---|---|---|---|---|---|
| STRIDE-8 | Testing set of derivation cohort | 8.8% | 85.0%<br>[84.5%–85.5%] | 89.6%<br>[89.3%–89.8%] | 33.4%<br>[32.3%–34.4%] | 99.0%<br>[98.9%–99.0%] | 89.3%<br>[89.0%–89.6%] | 96,112 / 191,426<br>1,650,202 / 16,968 |
| | Physician-adjudicated cohort | 8.8% | 86.1%<br>[83.5%–88.9%] | 76.2%<br>[70.1%–81.7%] | 51.9%<br>[44.3%–58.6%] | 94.9%<br>[93.5%–96.3%] | 78.5%<br>[73.9%–82.6%] | 2,432 / 2,258<br>7,236 / 391 |
| STRIDE-24 | Testing set of derivation cohort | 7.9% | 85.0%<br>[84.5%–85.6%] | 89.6%<br>[89.3%–89.9%] | 34.1%<br>[32.9%–35.1%] | 99.0%<br>[98.9%–99.0%] | 89.3%<br>[89.0%–89.6%] | 87,631 / 169,500<br>1,464,100 / 15,457 |
| | Physician-adjudicated cohort | 7.9% | 85.9%<br>[82.3%–89.1%] | 75.7%<br>[69.7%–81.0%] | 51.4%<br>[43.8%–58.5%] | 94.7%<br>[93.0%–96.1%] | 78.1%<br>[73.4%–82.3%] | 2,282 / 2,159<br>6,741 / 375 |
| STRIDE-48 | Testing set of derivation cohort | 6.0% | 85.0%<br>[84.3%–85.6%] | 89.0%<br>[88.7%–89.3%] | 31.7%<br>[30.4%–33.0%] | 99.0%<br>[98.9%–99.1%] | 88.8%<br>[88.5%–89.1%] | 67,868 / 145,997<br>1,183,820 / 11,971 |
| | Physician-adjudicated cohort | 6.0% | 86.2%<br>[82.1%–89.7%] | 74.3%<br>[67.7%–79.9%] | 47.4%<br>[39.7%–55.0%] | 95.2%<br>[93.5%–96.6%] | 76.8%<br>[71.5%–81.2%] | 1,905 / 2,111<br>6,096 / 305 |

Supplementary Table S9: STRIDE performance at 90% target sensitivity across observation windows. Thresholds selected on held-out derivation data as in the main-text operating-point table; means over five patient-level splits with 95% confidence intervals from a patient-level bootstrap. PPV, positive predictive value; NPV, negative predictive value; TP, FP, TN and FN, true positives, false positives, true negatives and false negatives (classified intervals, means per split).

| Method | Cohort | Threshold | Sensitivity | Specificity | PPV | NPV | Accuracy | TP / FP<br>TN / FN |
|---|---|---|---|---|---|---|---|---|
| STRIDE-8 | Testing set of derivation cohort | 6.3% | 90.0%<br>[89.6%–90.4%] | 87.8%<br>[87.5%–88.1%] | 31.3%<br>[30.1%–32.2%] | 99.3%<br>[99.3%–99.3%] | 88.0%<br>[87.6%–88.2%] | 101,762 / 223,820<br>1,617,807 / 11,318 |
| | Physician-adjudicated cohort | 6.3% | 91.3%<br>[89.3%–93.4%] | 74.2%<br>[67.8%–79.8%] | 51.3%<br>[44.1%–57.7%] | 96.6%<br>[95.6%–97.7%] | 78.1%<br>[73.4%–82.3%] | 2,578 / 2,451<br>7,043 / 245 |
| STRIDE-24 | Testing set of derivation cohort | 5.6% | 90.0%<br>[89.6%–90.4%] | 87.8%<br>[87.4%–88.1%] | 31.8%<br>[30.7%–32.8%] | 99.3%<br>[99.2%–99.3%] | 87.9%<br>[87.6%–88.2%] | 92,776 / 199,236<br>1,434,363 / 10,311 |
| | Physician-adjudicated cohort | 5.6% | 90.9%<br>[88.4%–92.9%] | 73.3%<br>[67.2%–79.0%] | 50.4%<br>[43.2%–57.5%] | 96.4%<br>[95.2%–97.5%] | 77.4%<br>[72.7%–81.6%] | 2,415 / 2,374<br>6,526 / 242 |
| STRIDE-48 | Testing set of derivation cohort | 4.1% | 90.0%<br>[89.4%–90.5%] | 87.2%<br>[86.9%–87.6%] | 29.8%<br>[28.5%–31.1%] | 99.3%<br>[99.3%–99.4%] | 87.4%<br>[87.1%–87.7%] | 71,856 / 169,558<br>1,160,259 / 7,983 |
| | Physician-adjudicated cohort | 4.1% | 91.0%<br>[87.9%–93.4%] | 72.1%<br>[65.5%–77.8%] | 46.8%<br>[39.3%–54.2%] | 96.8%<br>[95.4%–97.8%] | 76.2%<br>[70.9%–80.6%] | 2,012 / 2,286<br>5,921 / 198 |

Supplementary Table S10: ROC-AUC with and without linear interpolation of predictor values (means over five patient-level splits). Population, partitions and labels are identical in both analyses.

| **Method** | **Cohort** | **8-Hour** | | **24-Hour** | | **48-Hour** | |
|---|---|---|---|---|---|---|---|
| | | **With** | **Without** | **With** | **Without** | **With** | **Without** |
| STRIDE | Testing set of derivation cohort | 0.949 | 0.922 | 0.949 | 0.939 | 0.945 | 0.935 |
| | Physician-adjudicated cohort | 0.871 | 0.844 | 0.870 | 0.856 | 0.861 | 0.848 |
| SIRS | Testing set of derivation cohort | 0.891 | 0.891 | 0.886 | 0.886 | 0.886 | 0.886 |
| | Physician-adjudicated cohort | 0.848 | 0.848 | 0.840 | 0.840 | 0.837 | 0.837 |
| SOFA | Testing set of derivation cohort | 0.742 | 0.710 | 0.744 | 0.718 | 0.750 | 0.727 |
| | Physician-adjudicated cohort | 0.637 | 0.666 | 0.635 | 0.668 | 0.644 | 0.674 |
| Epic | Testing set of derivation cohort | 0.828 | 0.828 | 0.822 | 0.822 | 0.805 | 0.805 |
| | Physician-adjudicated cohort | 0.724 | 0.724 | 0.720 | 0.720 | 0.717 | 0.717 |

Supplementary Table S11: Discrimination of STRIDE-8 by hospital, calendar year and demographic group in the testing set of the derivation cohort. ROC-AUC and average precision (AP) are means over five patient-level splits with 95% confidence intervals from a patient-level bootstrap; interval and encounter counts are means per split. Hospitals are anonymized and ordered by number of encounters; hospital was assigned from the encounter department recorded in the EHR. Calendar year refers to the 8-hour interval, so an encounter spanning two years contributes to both. AP depends on the proportion of positive intervals and is not directly comparable across strata with different prevalence. † Affiliated non-acute sites with no positive intervals.

| Stratum | Positive intervals | Negative intervals | Encounters | Prevalence | ROC-AUC | AP |
|---|---|---|---|---|---|---|
| **All test intervals** | 113,081 | 1,841,627 | 101,535 | 5.8% | 0.949 [0.947–0.950] | 0.458 [0.445–0.470] |
| **Hospital** | | | | | | |
| Hospital A | 53,017 | 907,037 | 45,226 | 5.5% | 0.938 [0.935–0.941] | 0.407 [0.388–0.425] |
| Hospital B | 20,150 | 261,837 | 15,121 | 7.1% | 0.957 [0.954–0.960] | 0.544 [0.516–0.569] |
| Hospital C | 12,628 | 169,363 | 11,113 | 6.9% | 0.955 [0.952–0.958] | 0.524 [0.495–0.550] |
| Hospital D | 9,271 | 212,922 | 10,553 | 4.2% | 0.956 [0.952–0.960] | 0.416 [0.381–0.447] |
| Hospital E | 10,018 | 155,173 | 9,912 | 6.1% | 0.956 [0.953–0.959] | 0.510 [0.481–0.541] |
| Hospital F | 6,179 | 88,836 | 6,197 | 6.5% | 0.952 [0.948–0.956] | 0.494 [0.450–0.538] |
| Hospital G | 1,817 | 39,312 | 2,898 | 4.4% | 0.967 [0.962–0.971] | 0.518 [0.471–0.567] |
| Other sites† | 0 | 7,147 | 516 | 0.0% | – | – |
| **Year** | | | | | | |
| 2019 | 13,746 | 201,674 | 11,966 | 6.4% | 0.952 [0.947–0.956] | 0.505 [0.480–0.529] |
| 2020 | 16,523 | 220,998 | 12,478 | 7.0% | 0.943 [0.938–0.947] | 0.465 [0.436–0.491] |
| 2021 | 18,909 | 283,698 | 15,675 | 6.2% | 0.946 [0.941–0.950] | 0.453 [0.419–0.484] |
| 2022 | 18,609 | 303,182 | 16,287 | 5.8% | 0.948 [0.944–0.951] | 0.451 [0.425–0.476] |
| 2023 | 18,256 | 313,861 | 17,419 | 5.5% | 0.950 [0.947–0.953] | 0.457 [0.433–0.478] |
| 2024 | 18,250 | 334,527 | 18,749 | 5.2% | 0.951 [0.947–0.955] | 0.444 [0.414–0.475] |
| 2025 | 8,788 | 183,688 | 10,637 | 4.6% | 0.952 [0.948–0.956] | 0.436 [0.406–0.468] |
| **Sex** | | | | | | |
| Female | 48,423 | 893,440 | 52,523 | 5.1% | 0.952 [0.951–0.954] | 0.454 [0.438–0.468] |
| Male | 64,125 | 939,773 | 48,527 | 6.4% | 0.945 [0.943–0.947] | 0.461 [0.442–0.477] |
| Unknown | 532 | 8,415 | 485 | 5.9% | 0.951 [0.930–0.967] | 0.495 [0.300–0.629] |
| **Age band** | | | | | | |
| <50 years | 21,079 | 382,425 | 23,831 | 5.2% | 0.950 [0.946–0.953] | 0.439 [0.411–0.465] |
| 50–64 years | 31,598 | 452,054 | 24,212 | 6.5% | 0.945 [0.941–0.948] | 0.466 [0.443–0.490] |
| 65–79 years | 40,617 | 609,094 | 31,026 | 6.3% | 0.947 [0.944–0.950] | 0.470 [0.452–0.487] |
| ≥80 years | 19,267 | 389,947 | 22,003 | 4.7% | 0.954 [0.951–0.957] | 0.443 [0.420–0.464] |
| Unknown | 520 | 8,107 | 462 | 6.0% | 0.950 [0.927–0.966] | 0.490 [0.294–0.630] |
| **Race** | | | | | | |
| White | 81,820 | 1,328,851 | 73,506 | 5.8% | 0.950 [0.949–0.952] | 0.467 [0.453–0.481] |
| Black or African American | 11,939 | 209,532 | 10,848 | 5.4% | 0.945 [0.938–0.950] | 0.420 [0.380–0.453] |
| Other | 17,444 | 274,752 | 15,688 | 6.0% | 0.946 [0.942–0.950] | 0.450 [0.422–0.479] |
| Unknown | 1,878 | 28,493 | 1,492 | 6.2% | 0.933 [0.918–0.947] | 0.406 [0.326–0.477] |
| **Ethnicity** | | | | | | |
| Not Hispanic or Latino | 94,975 | 1,556,557 | 85,329 | 5.8% | 0.949 [0.947–0.951] | 0.459 [0.445–0.473] |
| Hispanic or Latino | 15,682 | 249,945 | 14,347 | 5.9% | 0.948 [0.944–0.951] | 0.457 [0.426–0.487] |
| Unknown | 2,424 | 35,125 | 1,858 | 6.5% | 0.936 [0.923–0.948] | 0.430 [0.356–0.498] |

Supplementary Table S12: Agreement between Llama classifications and physician adjudication in the 349 initially sepsis-coded encounters used for prompt development. Proportions are given with 95% Wilson confidence intervals; the F1 confidence interval was obtained from 10,000 bootstrap resamples of encounters.

| | **Physician: septic** | **Physician: not septic** | **Total** |
|---|---|---|---|
| **Llama: septic** | 287 (TP) | 16 (FP) | 303 |
| **Llama: not septic** | 19 (FN) | 27 (TN) | 46 |
| **Total** | 306 | 43 | 349 |

| **Measure** | **Value** | **95% CI** |
|---|---|---|
| Agreement (accuracy) | 90.0% (314/349) | 86.4–92.7% |
| Sensitivity | 93.8% (287/306) | 90.5–96.0% |
| Specificity | 62.8% (27/43) | 47.9–75.6% |
| Positive predictive value | 94.7% (287/303) | 91.6–96.7% |
| Negative predictive value | 58.7% (27/46) | 44.3–71.7% |
| F1 score | 0.943 | 0.922–0.961 |

Supplementary Table S13: STRIDE-8 discrimination under alternative allocations of Llama-reclassified encounters. Means over five patient-level splits (standard deviation across splits ≤0.0006 for derivation-test and ≤0.0043 for physician-adjudicated ROC-AUC). In designs A and B, patients with reclassified encounters could be allocated to testing, so the test sets differ from those of the main analysis.

| **Design** | **Test ROC-AUC** | **Test AP** | **Adjudicated ROC-AUC** | **Adjudicated AP** |
|---|---|---|---|---|
| Main analysis: reclassified patients in training only | 0.949 | 0.458 | 0.871 | 0.557 |
| A: reclassified patients split; reclassified encounters negative | 0.943 | 0.429 | 0.872 | 0.560 |
| A, test excluding reclassified encounters | 0.948 | – | – | – |
| A, reclassified encounters scored with original labels | 0.946 | 0.482 | – | – |
| B: reclassified encounters removed | 0.949 | 0.476 | 0.872 | 0.558 |

Supplementary Table S14: Electronic health record sources of the clinical variables. Each STRIDE variable is summarized as the minimum, mean and maximum within each 8-hour interval; urine output is the interval total. Flowsheet values are nursing and device documentation; laboratory values are assigned to intervals by their order date and time. Variables marked SOFA were used only for the SOFA comparator.

| **Variable** | **Source** | **EHR item(s)** | **Unit** |
|---|---|---|---|
| **Vital signs (STRIDE predictors)** | | | |
| Heart rate | Flowsheet | Pulse | beats/min |
| Respiratory rate | Flowsheet | Respirations | breaths/min |
| Systolic and diastolic blood pressure | Flowsheet | Blood pressure (systolic and diastolic components) | mmHg |
| Temperature | Flowsheet | Temperature; anesthesia temperature 1 and 2; Temperature (Celsius)[a] | °F |
| Oxygen saturation | Flowsheet | Pulse oximetry ($SpO_2$) | % |
| **Laboratory values (STRIDE predictors)** | | | |
| Albumin | Laboratory | Albumin (three serum components); | g/dL |
| Alanine aminotransferase | Laboratory | ALT | U/L |
| Alkaline phosphatase | Laboratory | Alkaline phosphatase | U/L |
| Aspartate aminotransferase | Laboratory | AST | U/L |
| Total bilirubin | Laboratory | Bilirubin, total | mg/dL |
| Blood urea nitrogen | Laboratory | BUN | mg/dL |
| Chloride | Laboratory | Chloride | mmol/L |
| $CO_2$ | Laboratory | $CO_2$ | mmol/L |
| Creatinine | Laboratory | Creatinine | mg/dL |
| Platelet count | Laboratory | Platelet count | $\times 10^9$/L |
| Potassium | Laboratory | Potassium | mmol/L |
| Sodium | Laboratory | Sodium | mmol/L |
| White blood cell count | Laboratory | White blood cell count | $\times 10^9$/L |
| **SOFA comparator only** | | | |
| $PaO_2$ | Laboratory | $pO_2$, arterial; $pO_2$ arterial, point of care | mmHg |
| $FiO_2$ | Flowsheet | Oxygen concentration (ventilator and ECMO documentation) | % |
| Mean arterial pressure | Flowsheet | Non-invasive mean arterial pressure (anesthesia record) | mmHg |
| Glasgow Coma Scale | Flowsheet | Glasgow Coma Scale score | points |
| Urine output | Flowsheet | Urine output (interval total) | mL |
| Weight | Flowsheet | Weight/scale[b] | kg |
| Lactate | Laboratory | Lactic acid (imputed, not scored) | mmol/L |
| Vasopressors | Medication administration | Adrenaline, noradrenaline, dopamine, dobutamine | $\mu$g/kg/min |

[a]Converted to °F. [b]Recorded in ounces and converted to kg.

Supplementary Table S15: Characteristics of included and excluded encounters. Excluded encounters failed at least one eligibility criterion (at least 48 hours of recorded data, no more than 40% missing values after interpolation, continuity, and, for sepsis-coded encounters, the operational clinical criteria). No age restriction was applied. Included encounters are the eligible cohort before LLM-based label refinement (Fig. 2). The original list of 614,346 screened non-sepsis-coded encounters was not retained. It was rebuilt by rerunning the original preprocessing on the July 2025 EHR extract, which recovered 610,540 encounters, and adding 2,529 eligible encounters preserved from the original cohort, giving 613,069 encounters for this comparison (286,310 included and 326,759 excluded); the remaining 1,277 could not be recovered. This reconstruction was used only for this comparison and does not affect the study cohort, the flow-diagram counts or any other analysis. In-hospital death was defined by a recorded death date during the admission or a discharge disposition of death. SIRS status could not be determined for excluded encounters because most lacked the required measurements.

| | **Sepsis-coded** | | **Non-sepsis-coded** | |
|---|---|---|---|---|
| **Characteristic** | **Included** | **Excluded** | **Included** | **Excluded** |
| Encounters | 66,105 | 11,289 | 286,310 | 326,759 |
| Age, mean (SD), years | 67.1 (16.8) | 68.4 (18.2) | 62.8 (19.7) | 41.2 (27.0) |
| Men, % | 52.8 | 50.3 | 47.2 | 42.0 |
| Length of stay, median (IQR), days | 6 (4–10) | 3 (1–4) | 4 (2–6) | 2 (1–3) |
| Length of stay 0–2 days, % | 10.2 | 48.5 | 25.1 | 69.5 |
| In-hospital death, % | 11.5 | 14.6 | 2.4 | 1.7 |
| Admission type, % | | | | |
| Emergency | 92.2 | 85.4 | 77.0 | 38.7 |
| Urgent | 5.1 | 10.7 | 6.5 | 10.8 |
| Elective | 1.5 | 3.3 | 13.1 | 33.5 |
| Other | 1.1 | 0.6 | 3.3 | 17.1 |

Supplementary Table S16: ICD-10-CM codes defining the sepsis-coded encounters. An encounter was sepsis-coded if a diagnosis recorded during the encounter, or its primary coded diagnosis, was an EHR diagnosis named as sepsis; each such diagnosis was mapped to ICD-10-CM through the EHR diagnosis terminology. Counts are encounters with at least one qualifying diagnosis mapped to the code; an encounter can carry several codes, so percentages do not sum to 100%.

| **Code** | **Description** | **Septic, derivation (n = 57,301)** | **Reclassified by Llama (n = 8,395)** | **Septic, adjudicated (n = 306)** | **Non-septic, adjudicated (n = 43)** |
|---|---|---|---|---|---|
| A41.9 | Sepsis, unspecified organism | 52,002 (90.8%) | 7,944 (94.6%) | 281 (91.8%) | 43 (100.0%) |
| A41.51 | Sepsis due to *Escherichia coli* | 5,523 (9.6%) | 184 (2.2%) | 37 (12.1%) | – |
| A41.89 | Other specified sepsis | 4,345 (7.6%) | 458 (5.5%) | 19 (6.2%) | 1 (2.3%) |
| A41.59 | Other gram-negative sepsis | 2,336 (4.1%) | 81 (1.0%) | 20 (6.5%) | – |
| R65.20 | Severe sepsis without septic shock | 2,225 (3.9%) | 220 (2.6%) | 9 (2.9%) | – |
| A41.01 | Sepsis due to methicillin susceptible *Staphylococcus aureus* | 2,174 (3.8%) | 94 (1.1%) | 12 (3.9%) | 1 (2.3%) |
| A41.02 | Sepsis due to methicillin resistant *Staphylococcus aureus* | 1,336 (2.3%) | 64 (0.8%) | 8 (2.6%) | – |
| A41.81 | Sepsis due to *Enterococcus* | 1,253 (2.2%) | 47 (0.6%) | 10 (3.3%) | – |
| A41.50 | Gram-negative sepsis, unspecified | 1,203 (2.1%) | 81 (1.0%) | 9 (2.9%) | – |
| A41.52 | Sepsis due to *Pseudomonas* | 1,154 (2.0%) | 49 (0.6%) | 6 (2.0%) | – |
| A41.4 | Sepsis due to anaerobes | 737 (1.3%) | 38 (0.5%) | 2 (0.7%) | – |
| A40.1 | Sepsis due to *Streptococcus*, group B | 610 (1.1%) | 20 (0.2%) | 3 (1.0%) | – |
| T81.44XA | Sepsis following a procedure, initial encounter | 593 (1.0%) | 53 (0.6%) | 4 (1.3%) | – |
| A40.3 | Sepsis due to *Streptococcus pneumoniae* | 492 (0.9%) | 23 (0.3%) | 7 (2.3%) | – |
| A40.9 | Streptococcal sepsis, unspecified | 478 (0.8%) | 21 (0.3%) | – | – |
| A40.8 | Other streptococcal sepsis | 473 (0.8%) | 13 (0.2%) | 4 (1.3%) | – |
| A40.0 | Sepsis due to *Streptococcus*, group A | 397 (0.7%) | 13 (0.2%) | 2 (0.7%) | – |
| A41.1 | Sepsis due to other specified *Staphylococcus* | 342 (0.6%) | 14 (0.2%) | – | 1 (2.3%) |
| R65.21 | Severe sepsis with septic shock | 327 (0.6%) | 25 (0.3%) | 1 (0.3%) | – |
| A41.53 | Sepsis due to *Serratia* | 265 (0.5%) | 9 (0.1%) | – | – |
| A41.2 | Sepsis due to unspecified *Staphylococcus* | 217 (0.4%) | 12 (0.1%) | 1 (0.3%) | – |
| A41.3 | Sepsis due to *Hemophilus influenzae* | 115 (0.2%) | 9 (0.1%) | – | – |
| B37.7 | Candidal sepsis | 94 (0.2%) | 2 (0.0%) | – | – |
| A02.1 | Salmonella sepsis | 87 (0.2%) | 9 (0.1%) | – | – |
| O85 | Puerperal sepsis | 40 (0.1%) | – | – | – |
| O86.04 | Sepsis following an obstetrical procedure | 25 (0.0%) | 4 (0.0%) | – | – |
| A41.54 | Sepsis due to *Acinetobacter baumannii* | 16 (0.0%) | 2 (0.0%) | – | – |
| O03.87 | Sepsis following complete or unspecified spontaneous abortion | 14 (0.0%) | – | – | – |
| A42.7 | Actinomycotic sepsis | 13 (0.0%) | – | – | – |
| O04.87 | Sepsis following (induced) termination of pregnancy | 12 (0.0%) | – | – | – |
| A32.7 | Listerial sepsis | 9 (0.0%) | – | – | – |
| P36.4 | Sepsis of newborn due to *Escherichia coli* | 8 (0.0%) | – | – | – |
| O03.37 | Sepsis following incomplete spontaneous abortion | 7 (0.0%) | – | – | – |
| O07.37 | Sepsis following failed attempted termination of pregnancy | 4 (0.0%) | – | – | – |
| T81.44XS | Sepsis following a procedure, sequela | 4 (0.0%) | – | – | – |

| Code | Description | Septic, derivation (n = 57,301) | Reclassified by Llama (n = 8,395) | Septic, adjudicated (n = 306) | Non-septic, adjudicated (n = 43) |
|---|---|---|---|---|---|
| P36.0 | Sepsis of newborn due to *Streptococcus*, group b | 3 (0.0%) | – | – | – |
| T81.44XD | Sepsis following a procedure, subsequent encounter | 3 (0.0%) | – | – | – |
| O08.82 | Sepsis following ectopic and molar pregnancy | 2 (0.0%) | – | – | – |
| P36.19 | Sepsis of newborn due to other streptococci | 2 (0.0%) | – | – | – |
| A26.7 | Erysipelothrix sepsis | 1 (0.0%) | – | – | – |
| P36.30 | Sepsis of newborn due to unspecified staphylococci | 1 (0.0%) | – | – | – |

Supplementary Table S17: ROC-AUC of the full and SIRS-feature-ablated STRIDE-8 models in the derivation-test comparison groups. The ablated model was retrained without the minimum, mean and maximum temperature, heart rate, respiratory rate and white blood cell count, using the same outcome labels and patient partitions. Values are means over five patient-level splits.

| Comparison group | STRIDE-8, full model | STRIDE-8, without SIRS features |
|---|---|---|
| Septic subset | 0.907 | 0.755 |
| Septic + non-septic SIRS+ encounters | 0.926 | 0.819 |
| Septic + non-septic SIRS− encounters | 0.963 | 0.861 |
